# Carriers of *LRRK2* pathogenic variants show a milder, anatomically distinct brain signature of Parkinson’s disease

**DOI:** 10.1101/2025.03.09.25323610

**Authors:** Jakub Kopal, Andrew Vo, Qin Tao, Tanya Simuni, Lana M. Chahine, Danilo Bzdok, Alain Dagher

## Abstract

*LRRK2* gene variants are a major genetic risk factor for both familial and sporadic Parkinson’s disease (PD), opening an unattended window on the disease’s mechanisms and potential therapies. Investigating the influence of pathogenic variants in *LRRK2* gene on brain structure is a crucial step toward enabling early diagnosis and personalized treatment. Yet, despite its significance, the ways in which *LRRK2* genotype affects brain structure remain largely unexplored. Work in this domain is plagued by small sample sizes and differences in cohort composition, which can obscure genuine distinctions among clinical subgroups. In this study, we overcome such important limitations by combining explicit modeling of population background variation and pattern matching. Specifically, we leveraged a large cohort of 641 participants (including 364 with a PD diagnosis) to examine MRI-detectable cortical atrophy patterns associated with the *LRRK2* pathogenic variants in people with PD and non-manifesting individuals. LRRK2 PD patients exhibited milder cortical thinning compared to sporadic PD, with notable preservation in temporal and occipital regions, suggesting a distinct pattern of neurodegeneration. Non-manifesting LRRK2 carriers showed no significant cortical atrophy, indicating no structural signs of subclinical PD. We further analyzed the relationship between aggregated alpha-synuclein in cerebrospinal fluid and atrophy. We found that those with evidence of aggregated alpha-synuclein experienced pronounced neurodegeneration and increased cortical thinning, possibly defining another aggressive PD subtype. Our findings highlight avenues for distinguishing PD subtypes, which could lead to more targeted treatment approaches and a more complete understanding of Parkinson’s disease progression.

## Introduction

Monogenic genetic Parkinson’s Disease (PD) is rare, but studying disease-associated pathogenic variants has helped identify biological processes generally involved in sporadic PD. Mutations in the gene coding for Leucine-Rich Repeat Kinase 2 (*LRRK2*) are the commonest cause of familial PD^1,2^. The inheritance is autosomal dominant with age-dependent penetrance, from 28% at age 59 to 74% at 79 years^3^. *LRRK2* has emerged as a potential therapeutic target in both LRRK2-associated but also sporadic PD (sPD). Prevalence of *LRRK2* pathogenic variants among PD individuals is estimated to be 1-5% and is substantially higher in certain ethnic subgroups^4^. More importantly, although LRRK2 PD is relatively infrequent, there is post-mortem evidence of upregulation of *LRRK2* activity in the brains of people with sPD^5,6^. Because the disease-causing allele is a toxic gain of function mutation, *LRRK2* may be an attractive therapeutic target for inhibitory agents, which are already in clinical trials^1^. Indeed, growing evidence suggests that *LRRK2* kinase inhibitors may offer neuroprotective benefits in PD by restoring lysosomal function^7^. Therefore, gaining a deeper understanding of neurodegeneration in individuals with *LRRK2*- associated PD may help distinguish *LRRK2*-specific mechanisms from broader PD pathology, offering critical insights into disease heterogeneity, progression, and potential compensatory pathways. This could also aid in the development of precise imaging biomarkers for tracking *LRRK2*-targeted therapeutic responses and refining personalized treatment strategies for PD.

PD associated with *LRRK2 pathogenic* variants has a clinical picture similar to sPD. Patients develop levo-dopa responsive parkinsonism with a mean age of onset in the sixth decade^8^. They show evidence of dopamine neuron degeneration on single-photon emission computed tomography dopamine transporter imaging (DATscan) similar to sPD^9^. However, important differences have also been noted. LRRK2 PD appears to be milder with slower progression^1,10^. More importantly, only 40-75% of LRRK2 PD patients show characteristic Lewy pathology at autopsy^3^, suggesting that neurodegeneration in LRRK2 PD sometimes occurs in the absence of synucleinopathy. Of the roughly one third of patients without synucleinopathy at postmortem, some demonstrate Alzheimer-like pathology, while others have dopamine neuron loss without evidence or protein aggregation^3,11^.

While it is not currently possible to quantitatively measure central nervous system Lewy pathology in living humans, amplification techniques similar to those used in prion diseases have been developed to detect aggregated alpha-synuclein (asyn) in cerebrospinal fluid (CSF)^12^. A positive CSF seed amplification assay (SAA) for alpha-synuclein indicates Lewy pathology with high sensitivity and specificity aside from amygdala predominant alpha- synuclein distribution^13,14^. In LRRK2 PD, CSF asyn SAA is congruent with postmortem examinations of substantia nigra synucleinopathy^15^ and may also detect evidence of synucleinopathy in non manifesting at-risk individuals^12^. Consistent with postmortem studies, CSF asyn SAA is positive in roughly 67% of LRRK2 PD individuals^12^. A negative CSF asyn SAA in LRRK2 PD appears to be associated with milder motor manifestations and slower progression^16^, positioning asyn SAA as an important disease biomarker.

Magnetic resonance imaging (MRI) serves as a crucial tool for differential diagnosis of PD and exploring its neuropathological mechanisms. The most useful MRI-derived indicators of neurodegeneration so far include cortical thickness (from T1-weighted MRI) and susceptibility-weighted or diffusion-weighted MRI of the substantia nigra. However, there have been few reports on the MRI-detectable characteristics of LRRK2 PD^9^. The studies published to date suffer from small sample sizes (typically below N = 20), where the evidence is further obscured by group differences in key sociodemographic and clinical factors. The Parkinson’s Progression Markers Initiative (PPMI) has collected MRI on an unprecedented number of sPD and LRRK2 PD with and without CSF asyn SAA positivity, as well as non-manifesting carriers (NMC)^17^. This extensive dataset opens a new window of opportunity to advance our understanding of PD through comprehensive MRI analysis. However, this cohort also faces limitations commonly seen in LRRK2 studies. Specifically, there are important demographic differences in PPMI between sPD and LRRK2 PD with respect to disease duration, subject age, age of onset, and scanning site, making direct comparisons challenging. Matching on all these variables would once again yield small sample sizes, while using all the data leaves one vulnerable to important confounds. Here, we propose a principled hierarchical approach with a propensity score matching analytical protocol to overcome these limitations. This pattern-matching study details cortical atrophy distribution in LRRK2 PD and NMC, and evaluates the influence of CSF asyn SAA status. We show that (1) it is possible to account for unmatched datasets and known confounds, (2) cortical and subcortical atrophy in LRRK2 PD is milder than sPD, with a distinct spatial pattern, and (3) asyn SAA status correlates with greater degree of tissue loss.

## Results

### Differences in key clinical variables within Parkinson’s disease dataset

We set out to unravel the effects of *LRRK2* pathogenic variants as well as the effect of asyn SAA status in a generously profiled cohort of participants with PD and controls (Table 1). This cohort of 641 participants consisted of participants with sporadic PD and no known monogenic variant (n=288), LRRK2 PD (n = 76), healthy controls (n = 133), and non-manifesting carriers of a *LRRK2* pathogenic variants (n = 94) (Fig. 1a). We further stratified our cohort based on CSF asyn SAA status. Across all participants, 295 displayed positive asyn SAA result and 149 were asyn SAA negative (Table 2). The asyn SAA was not available for the remaining participants.

**Figure 1:**
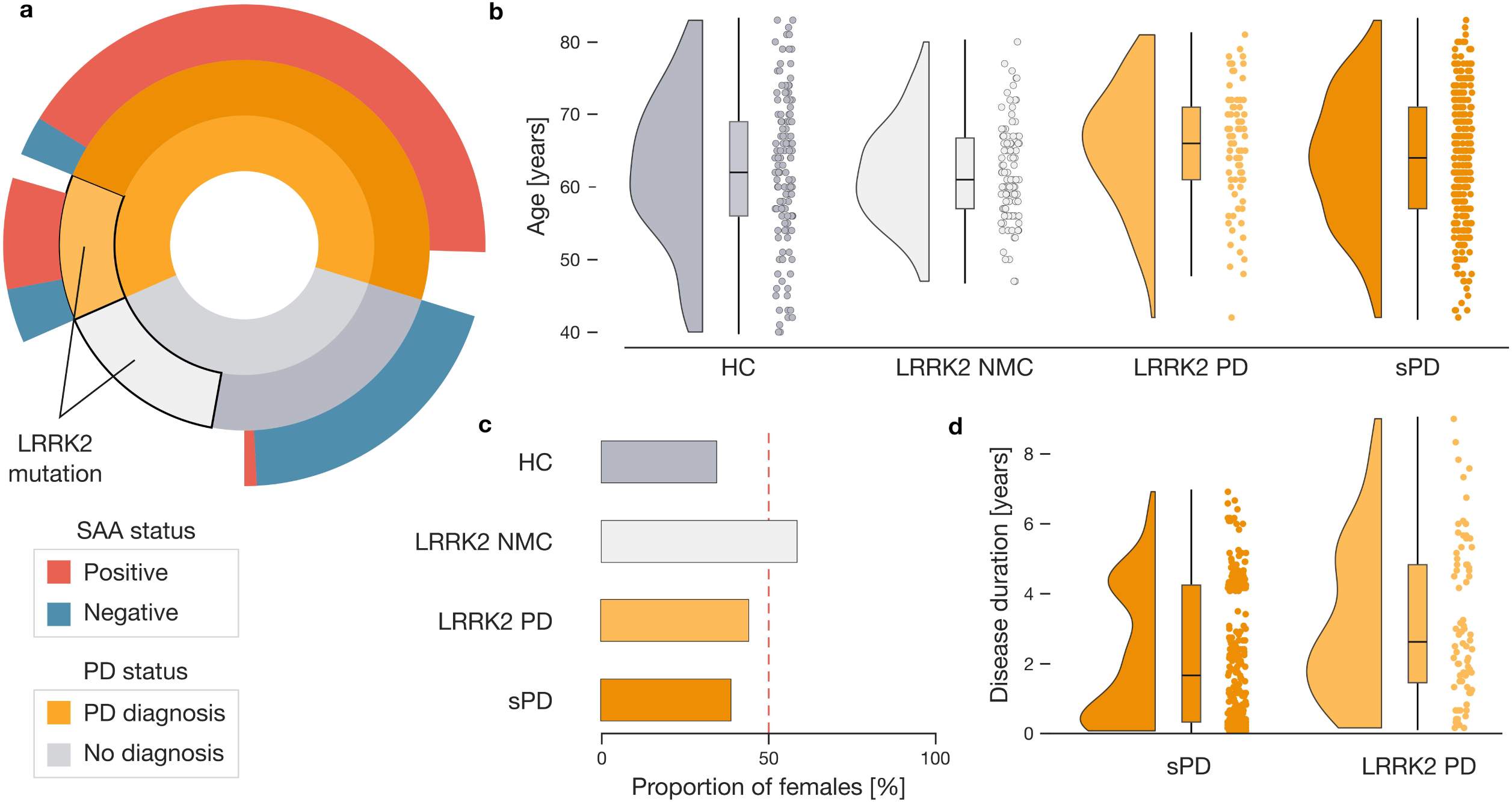
Complex interrelations between subgroups in ∼600 participant population cohort. **a.** Ratio breakdown of participant subgroups. Our participant sample consists of 364 participants with the diagnosis of Parkinson’s disease and 227 participants without PD diagnosis (inner circular bar plot). Among these, 94 non-PD participants and 76 PD patients carry *LRRK2* pathogenic variants indicated by the black edge (middle circular bar plot). Finally, the outer circular bar plot depicts the ratio of asyn SAA positive (red) and asyn SAA negative (blue) participants in each subgroup. **b.** PD patients carrying *LRRK2* pathogenic variants represent the oldest subgroup. Raincloud plots display the age of each participant in one of the four subgroups. **c.** Proportion of females among subgroups. Non-manifesting carriers with *LRRK2* pathogenic variants display the highest proportion of females. The red dotted line marks an equal ratio of male and female participants. **d.** Different disease duration in PD patients. Disease duration is plotted for PD patients separated by the presence of *LRRK2* pathogenic variant. The diversity in key demographic metrics poses a challenge to comparing PD and control subgroups. Abbreviations: HC=healthy controls, NMC=non-manifesting carrier, PD=Parkinson’s disease, sPD=sporadic PD.

**Table 1:**
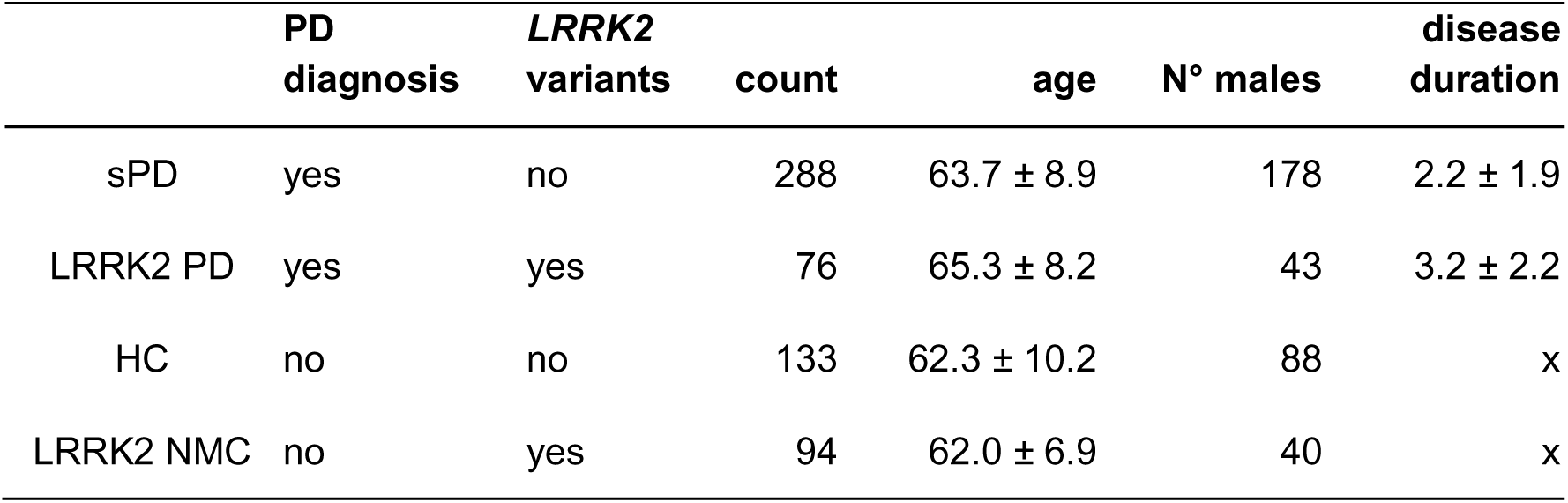
Dataset demographics, diagnosis, and LRRK2. Key participant characteristics are provided with respect to the diagnosis of Parkinson’s disease and the presence of *LRRK2* pathogenic variants. Age and disease duration are presented as mean ± standard deviation. Abbreviations: sPD=sporadic Parkinson’s disease, HC=healthy control, NCM=non-manifesting carrier.

|  | PD<br>diagnosis | *LRRK2*<br>variants | count | age | N° males | disease<br>duration |
| --- | --- | --- | --- | --- | --- | --- |
| sPD | yes | no | 288 | 63.7 ± 8.9 | 178 | 2.2 ± 1.9 |
| LRRK2 PD | yes | yes | 76 | 65.3 ± 8.2 | 43 | 3.2 ± 2.2 |
| HC | no | no | 133 | 62.3 ± 10.2 | 88 | x |
| LRRK2 NMC | no | yes | 94 | 62.0 ± 6.9 | 40 | x |

**Table 2:**
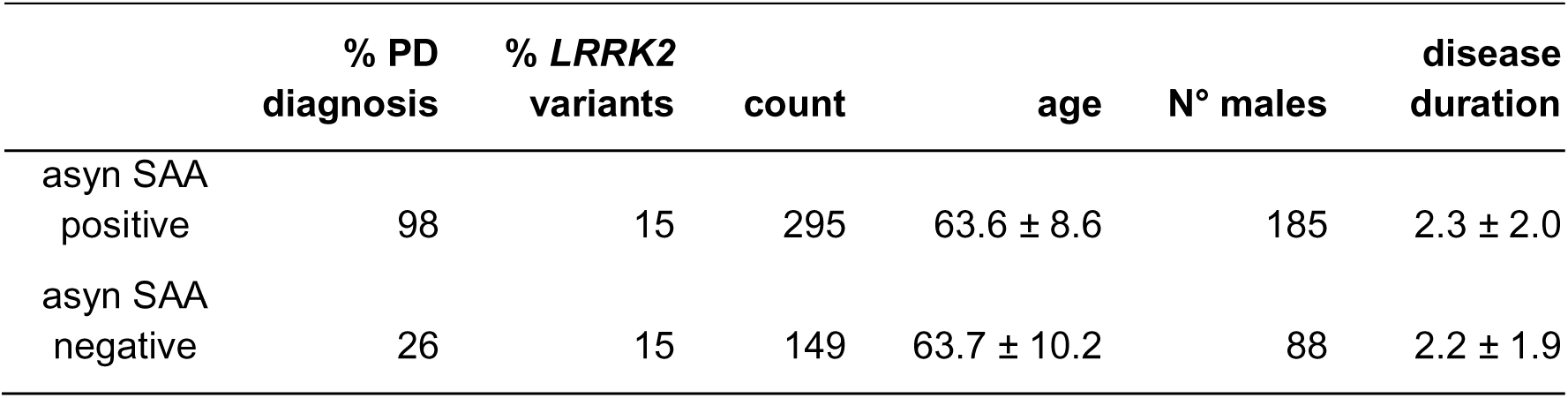
Dataset demographics and SAA status. Key participant characteristics are provided with respect to the asyn SAA status. Age and disease duration are presented as mean ± standard deviation. % PD diagnosis column corresponds to the percentage of asyn SAA positive participants that carry PD diagnosis.

|  | % PD<br>diagnosis | % *LRRK2*<br>variants | count | age | N° males | disease<br>duration |
| --- | --- | --- | --- | --- | --- | --- |
| asyn SAA<br>positive | 98 | 15 | 295 | 63.6 ± 8.6 | 185 | 2.3 ± 2.0 |
| asyn SAA<br>negative | 26 | 15 | 149 | 63.7 ± 10.2 | 88 | 2.2 ± 1.9 |

The hierarchical constellation of participant attributes, along with the existence of overlapping subgroups defined by disease, variant, and asyn SAA status, posed an analytic challenge due to the differences in key demographic parameters among the subgroups. Specifically, for the subgroups defined by disease and variant status, the average age in the LRRK2 PD subgroup was 65.3 ± 8.2 years, while in LRRK2 NMC it was 62.0 ± 6.9 years (Fig. 1b). In addition, the LRRK2 NMC subgroup contained the highest proportion of females (57 %), while healthy controls (HC) contained the lowest proportion (34 %) (Fig. 1c). Finally, we noticed a difference in disease duration between sPD and LRRK2 PD; 2.2 ± 1.9 years vs. 3.2 ± 2.2 years (Fig. 1d). Besides the differences among the four subgroups defined by PD diagnosis and *LRRK2* status, further demographic differences existed among subgroups defined by asyn SAA status (Sup. Fig. 1). Consequently, the complex relationship between key demographic factors called for a special approach to form homogenized and comparable subgroups for targeted analysis that is not unduly contaminated by unattended sources of general population variation^18,19^.

### Extracting a model of Parkinson’s disease brain atrophy

In order to overcome analytical challenges posed by the dataset structure, we first derived a quantitative model that explained PD as a function of structural brain alterations in carefully matched groups of sPD and HC participants (Fig. 2a). The matching strategy was based on propensity scores that considered age, sex, and site (cf. Methods). This approach guaranteed that PD diagnosis could not be predicted using these demographic factors alone. We thus selected 104 sPD and 104 HC matched participants for further analyses (Fig. 2b, c). For each brain region at hand, we estimated a separate linear regression model using PD diagnosis to predict regional thickness for cortical regions and regional volume for subcortical regions (Sup. Fig. 2). Examining associated explained variance (R^2^ , coefficient of determination) demonstrated that diagnosis accounted for a substantial portion of the variation in brain structure (Sup. Fig. 3). The majority of the estimated linear regression slope coefficients yielded negative effects, indicating cortical and subcortical atrophy associated with PD diagnosis (Fig. 2d). We observed strongest atrophy effects in temporal and occipital brain regions, namely right parahippocampal gyrus (ß = -0.37), banks of the superior temporal sulcus (ß = -0.30), and left precuneus (ß = -0.29). Notably, the obtained general pattern of PD-related atrophy mirrored previous structural brain-imaging studies^20^. The effects of PD diagnosis were further exacerbated for participants with higher age (Sup. Fig. 2). In summary, estimating a collection of dedicated linear models, applied to a carefully matched groups of sPD and HC participants, reliably captured expected, previously shown, structural alteration observed in PD patients.

**Figure 2:**
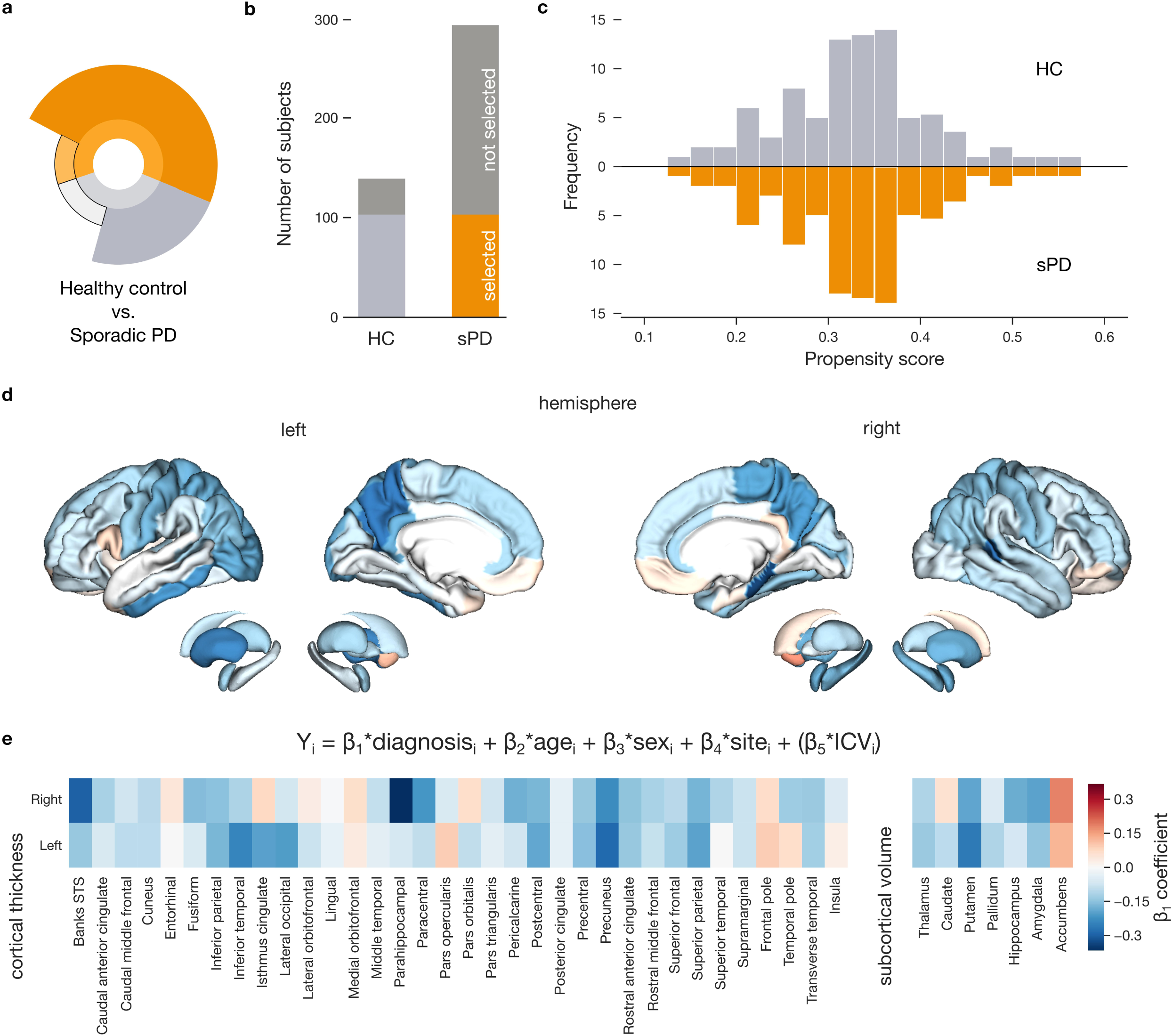
Region-specific models accurately estimate cortical thinning in PD. **a.** Subgroups used to derive models of general PD alterations. The infographic highlights the HC and sPD subgroups used for the ensuing analysis. **b.** Selection of carefully matched groups of participants. Only 104 out of 133 HC, resp. 104 out of 288 sPD, participants are carried forward to derive estimates of brain alterations associated with PD. **c.** Propensity scores enable careful participant matching. Participants from both subgroups are matched based on age, sex, and site using a flexible propensity score matching strategy, resulting in an excellent match. The histograms display frequencies of propensity scores separately for sPD and HC subgroups. **d.** Estimates of brain structure alterations. Regional thickness for cortical regions and regional volume for subcortical regions are predicted using a series of linear models based on disease diagnosis, age, sex, and site. Colored brains display the diagnosis coefficient estimates highlighting decreased (blue color) or increased (red color) thickness, resp. volume. **e.** Effect size of diagnosis across all brain regions. The heatmap shows diagnosis beta coefficients among 68 regions based on the Desikan-Killiany cortical parcellation and 16 regions defined by Harvard-Oxford subcortical atlas. Detected cortical thinning and volume loss aligns with established PD changes, underscoring our analytical protocol.

### Delineating neurodegeneration changes associated with *LRRK2* pathogenic variant

After estimating the PD-related atrophy in participants without *LRRK2* pathogenic variants, we directed our attention to the effects of these variants. To that end, we compared Parkinson’s patients carrying the *LRRK2* pathogenic variants against patients without this genetic status (Fig. 3a). We again carefully matched groups of sPD and LRRK2 PD participants using propensity scores taking into account age, sex, disease duration, and site. We thus selected 19 patients with and 19 without the variants (Fig. 3b). To address the low statistical power due to the small sample size, we applied models trained in earlier steps using a larger participant set in the new subgroups, thus avoiding the necessity of estimating new model parameters. In other words, we carried the model of general PD atrophy over to these new subgroups defined by the presence of the *LRRK2* pathogenic variants. Specifically, we derived regional morphometry (i.e., cortical thickness and subcortical volume) estimates for each sPD and LRRK2 PD patient using the linear models derived in HC vs. sPD comparison (i.e., in the previous step). We then compared how these regional estimates differed from actual brain measurements (Fig. 3d). Finally, after averaging the differences between morphometry estimates and the actual measurement across all participants in each subgroup, we obtained a single measure of prediction difference for each brain region for the considered participants - sPD and LRRK2 PD (Sup. Fig. 4). We observed a statistically significant difference between the two subgroups in these modeling outcomes (two-sided t-test p = 1.1 x 10^-6^). Notably in LRRK2 PD patients, the actual thickness and volumes were more preserved compared to those estimated by our model of PD atrophy pointing towards less cortical thinning and volume reduction in LRRK2 PD patients compared to sPD. In other words, our participant matching strategy combined with the transfer of PD atrophy model revealed significantly milder brain atrophy in patients with *LRRK2* pathogenic variants.

**Figure 3:**
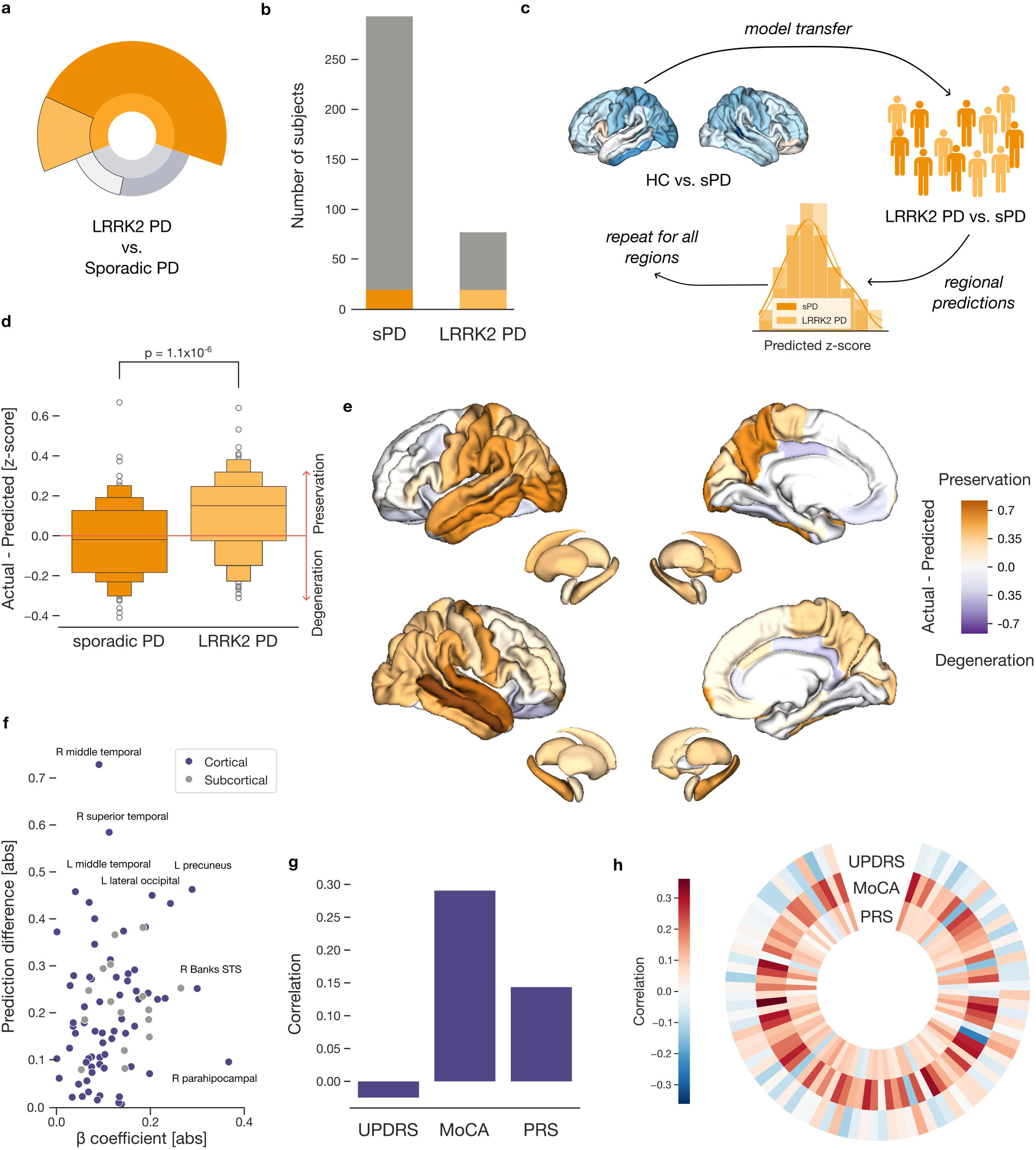
Parkinson’s patients with *LRRK2* pathogenic variants show a milder form of brain atrophy. **a.** Subgroups used to assess the extent of PD-related atrophy. The infographic indicates the sPD and LRRK2 PD subgroups used for the present analysis. **b.** Selection of carefully matched groups of PD patients. Only 19 out of 288 sPD, resp. 19 out of 76 LRRK2, participants are selected based on age, sex, disease duration and site using propensity score matching. **c.** Transferring the model of PD-related atrophy between cohort subgroups. We apply a PD atrophy model, trained on measures from sPD and HC subgroups, to estimate regional thickness and volume in LRRK2 PD and sPD patients (i.e., model transfer). We then average these estimates across participants to derive region-specific measures. **d.** Model estimates reveal less atrophy in LRRK2 PD patients. We compare region-wise differences of measured and estimated morphometry between the two subgroups of PD patients. Positives suggest preservation of brain structure while negatives point towards pronounced degeneration. The p-value denotes a two-sample t-test of prediction differences. **e.** *LRRK2* pathogenic variants are associated with less cortical thinning in temporal and occipital regions. Region-wise differences between measured and estimated brain metrics are plotted on cortical and subcortical regions for LRRK2 PD participants. Orange color highlights regions with lesser amount of cortical thinning or volume reduction while purple color denotes increased thinning or reduction. **f.** Relationship of prediction difference with PD atrophy. The diagnosis-related beta coefficients are plotted against prediction differences for every brain region. **g.** Linking predictions of brain structure to cognitive and genetic measures. The correlations quantify linear association strength across all sPD and LRRK2 PD patients between predictions of brain structure averaged across all regions with MDS-UPDRS III, MoCA, and PRS. **h.** Relating regional predictions to cognitive and genetic factors. Correlations between MDS-UPDRS III, MoCA, and PRS with predictions of regional morphometry are displayed for all regions. Overall, patients carrying *LRRK2* pathogenic variants show less brain atrophy, with preservation especially in the temporal and occipital regions.

We then aimed to investigate if the uncovered pattern of milder cortical thinning and volume reduction resembled the pattern of PD-related brain atrophy. Therefore, we first examined which brain regions exhibited the largest differences between measured and predicted morphometry in LRRK2 PD participants. Larger measured morphometry compared to predicted morphometry corresponded to milder cortical thinning and thus preservation. In general, regions with the highest preservation were located predominantly in the temporal and occipital lobe (Fig. 3e). Concretely, the largest preservations of thickness were observed in the left and right middle temporal gyrus, right superior temporal gyrus, or in the left precuneus.

To formally compare this pattern of preservation to the previously obtained pattern of PD-related brain atrophy (cf. above), we calculated Pearson’s correlation across the two sets of brain region measures (i.e., model coefficients and prediction differences mapped onto the brain). We used a spin-permutation test across the whole brain to examine the statistical significance of Pearson’s correlation between brain maps. The two brain maps were not statistically similar (r = 0.20, p_spin_ > 0.05) and especially regions such as right parahippocampal gyrus displayed large divergence between atrophy and preservation (Fig. 3f). In contrast, left precuneus and lateral occipital gyrus displayed substantial cortical thinning as well as preservation in LRRK2 PD patients. This suggests that the *LRRK2* pathogenic variants may be linked to a distinct spatial pattern of brain changes compared to typical sPD atrophy.

After establishing LRRK2 PD as a milder and distinct version of PD, we sought to explore how clinical severity of PD, individual differences in cognitive function, and genetic risk liability relate to brain structure predictions. Specifically, we examined how the Movement Disorders Society Unified Parkinson’s Disease Rating Scale (UPDRS) part III correlates with predicted measurements of expected brain morphometry to assess the impact of brain atrophy on motor manifestations. Furthermore, the relationship between the Montreal Cognitive Assessment (MoCA) scores and predicted morphometry provided insights into how global cognitive function in PD is linked to specific structural brain changes. Finally, associations with polygenic risk scores (PRS) for Parkinson’s disease allowed us to explore genetic risk to brain structure alterations in PD. In the first step, we used such predictions of brain thicknesses and volumes averaged across all regions for all LRRK2 PD and sPD patients (Fig. 3g). We observed significant Pearson’s correlation with MoCa, where decreased thickness was linked to lower cognitive scores (r_MoCA_ = 0.29, p_FDR_ = 0.001). The other two tests did not reach significance. Notably, these associations exceeded associations with actual measured structure (Sup. Fig. 5). Collectively, this significant association links previously observed better cognitive performance of LRRK2 PD compared to sPD subjects and their reduced cortical thinning.

In the second step, we explored the relationship between MDS-UPDRS part III, MoCA, and PRS with predicted morphometry of each region separately. From a broader perspective, the associations followed the anticipated directionality (Fig. 3h). The strongest link with cognitive impairment represented by MoCA scores was with the postcentral gyri (r_left_ = -0.38; r_right_ = 0.26), the paracentral gyri (r_left_ = 0.33; r_right_ = 0.33), and the superior temporal gyrus (r = 0.32). Meanwhile, the left caudal middle frontal gyrus (r = -0.15), and the right putamen (r = -0.12), displayed the strongest link to motor manifestations (UPDRS) (Sup. Tab. 2). Although the associations did not remain significant after FDR correction for the 84*3 tests, the uncovered patterns can guide future research efforts. In conclusion, these results demonstrated that LRRK2 PD patients exhibited considerably less cortical thinning and volume reduction compared to sPD patients, particularly in temporal and occipital regions. The observed association patterns between clinical severity, cognitive function, and brain structure offer valuable insights that may inform future research directions in understanding the nuances of Parkinson’s disease progression.

In addition to studying the effects of the LRRK2 pathogenic variants in PD patients, we also assessed the impact of the variants on participants without a PD diagnosis. Analogous to previous steps, we first selected carefully matched subgroups of HC and LRRK2 NMC participants (Fig. 4a). Propensity score matching based on age, sex, and site indicated 42 closely matched pairs of participants without PD diagnosis (Fig. 4b). We again leveraged the opportunity to transfer readily-deployable models from the HC vs. sPD comparison (Fig. 4c). Therefore, we calculated regional morphometry for each HC and LRRK2 NMC participant using the pre-estimated models. We then compared the obtained regional estimates to actual brain measurements and averaged the differences across all participants in each subgroup (Sup. Fig. 3). Based on these single region-specific estimates for HC and LRRK2 NMC subgroups, we did not observe a significant difference between the HC and LRRK2 NMC subgroups (two-sided t-test p = 0.59) (Fig. 4d). Our findings thus indicated that the *LRRK2* pathogenic variants do not profoundly affect brain structure in healthy participants without a PD diagnosis.

**Figure 4.**
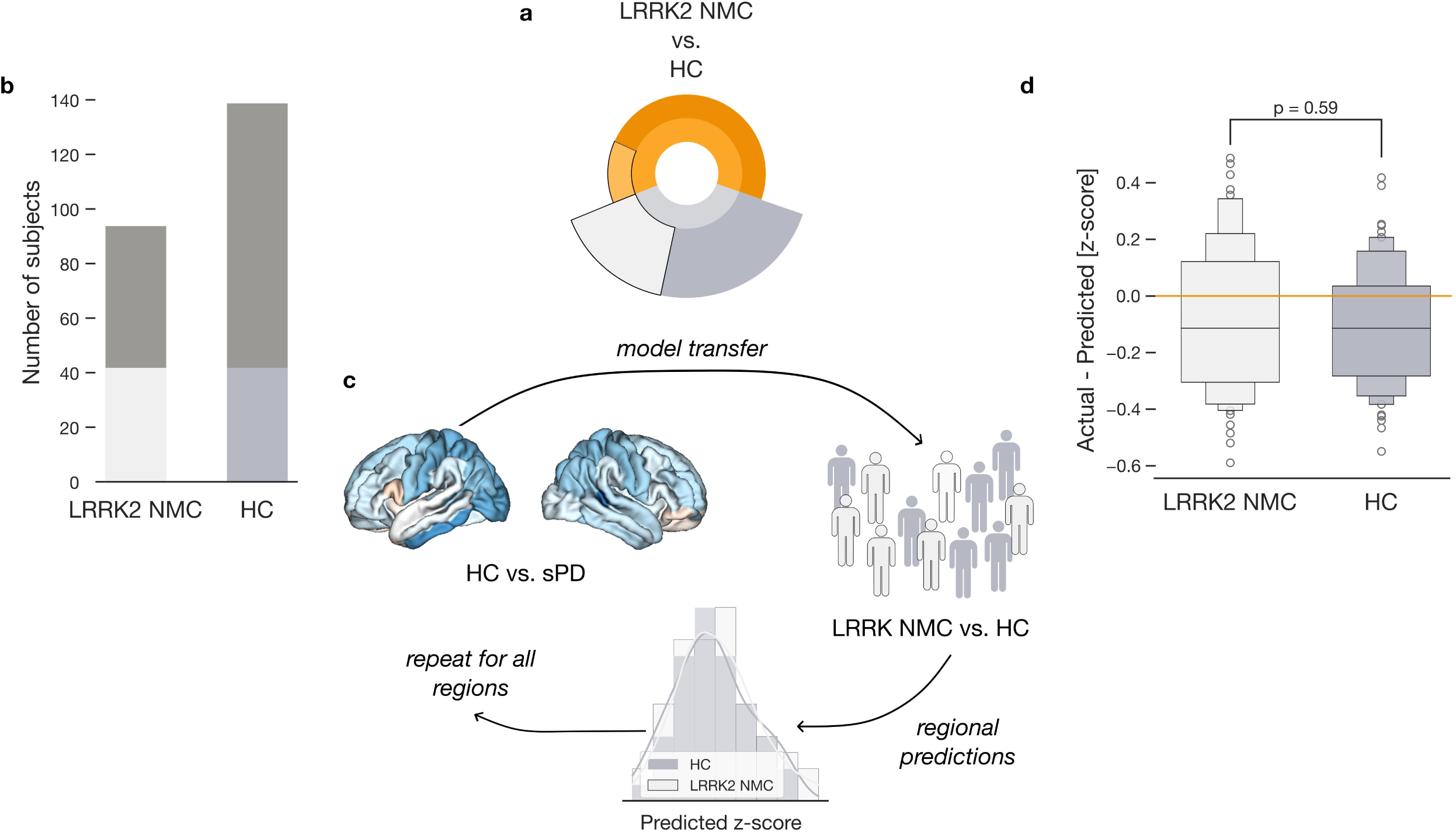
Absence of significant brain structure alterations in LRRK2 non-manifesting carriers. **a.** The influence of *LRRK2* pathogenic variants in participants without PD diagnosis. We analyze the influence of *LRRK2* pathogenic variants in HC and LRRK2 NMC participants. **b.** Selection of carefully matched groups of HC and LRRK2 NMC participants. Only 42 HC and 42 LRRK2 NMC participants are selected based on age, sex, and site using propensity score matching. **c.** Transferring the model of PD-related atrophy. We apply a PD atrophy model, trained on measures from sPD and HC subgroups, to estimate regional brain thickness and volume in HC and LRRK2 NMC. **d.** Comparable estimates of brain morphometry in controls. Boxen plots display region-wise differences of measured and estimated morphometry for HC and LRRK2 NMC subgroups. The p-value denotes a two- sample t-test of prediction differences. Unlike in PD patients, the presence of *LRRK2* pathogenic variants alone is not associated with significant structural brain alterations in participants without PD diagnosis.

### Relationship between aggregated alpha-synuclein and brain atrophy in Parkinson’s disease

Our final goal was to investigate the relationship between aggregated alpha-synuclein and PD-related brain atrophy. Repeating previously utilized methodology, from all available participants, we selected 23 asyn SAA positive participants, and 23 participants without that attribute, who we matched using propensity scores based on age, sex, site, and disease duration (Fig. 5a,b). Considering the small number of matching subjects, we transferred the model of PD-related atrophy to obtain an estimate of morphometry for each brain region in each matched participant (Sup. Fig. 3). These estimates enabled us to examine how aggregated asyn in CSF relates to PD-related brain atrophy in rigorously matched participants.

**Figure 5.**
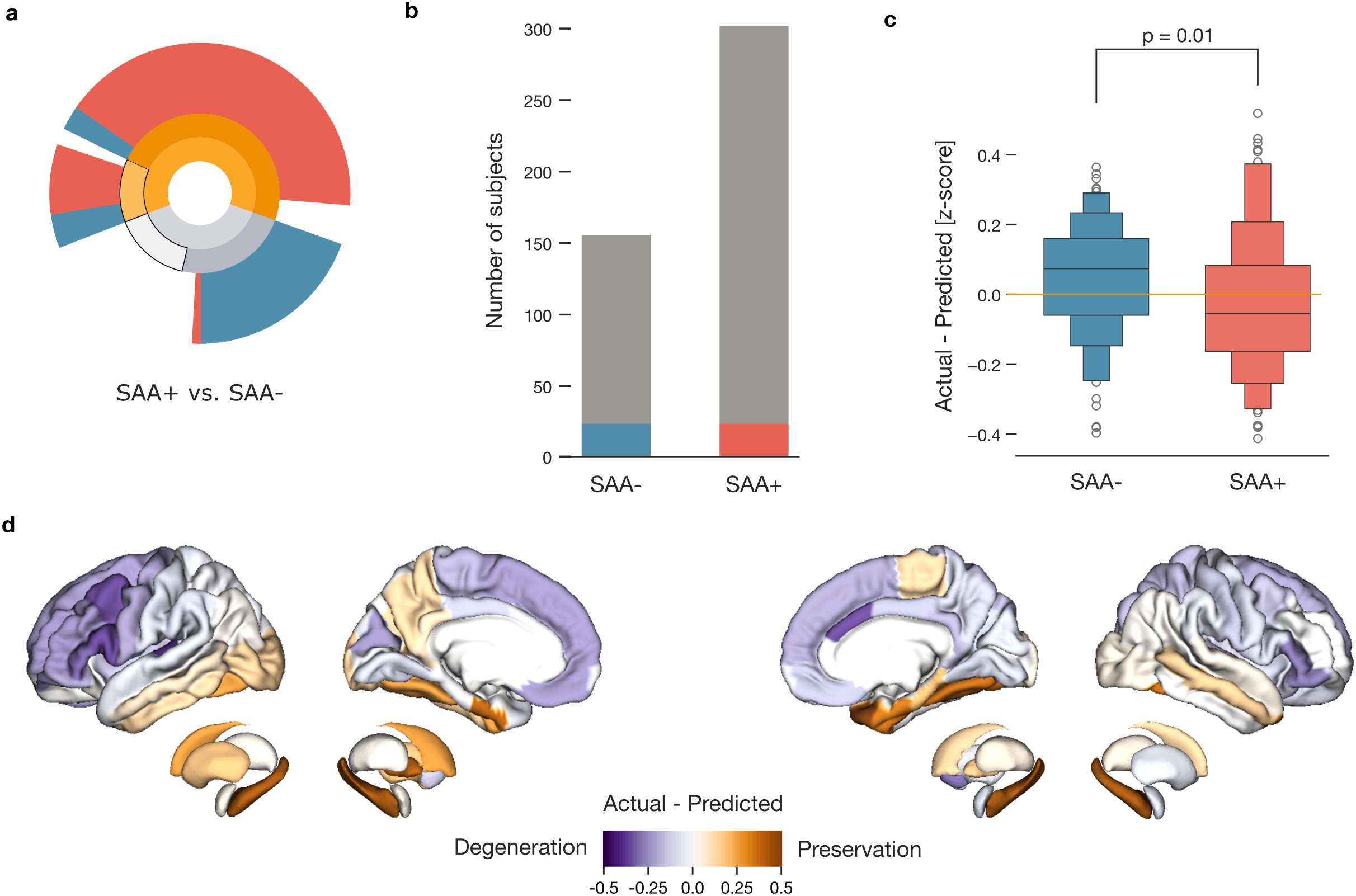
Aggregated alpha-synuclein proteins associated with increased brain atrophy. **a.** Measures of asyn SAA across participant subgroups. We analyze the influence of asyn SAA measured in HC, sPD, and LRRK2 PD participants. **b.** Only 23 out of 149 asyn SAA negative, resp. 23 out of 295 asyn SAA positive, participants are selected based on age, sex, disease duration and site using propensity score matching. **c.** Model estimates show more atrophy associated with asyn SAA positive status. We compare region-wise differences of measured and estimated morphometry between groups of asyn SAA positive and negative participants. Lower measured compared to predicted thickness and volume suggests pronounced degeneration. **d.** Differences in vulnerability for degeneration in cortical and subcortical regions associated with positive asyn SAA result. Region-wise differences between measured and estimated brain morphometry for asyn SAA positive are plotted on cortical and subcortical regions. Orange color highlights regions with lesser amount of cortical thinning or volume reduction than expected by the model PD atrophy while purple color denotes increased thinning or reduction. Our findings reveal pronounced cortical thinning specifically linked to asyn SAA positive status, with distinct cortical regions showing increased vulnerability.

Comparing obtained regional estimates to actual brain measurements revealed a significant difference between the asyn SAA positive and asyn SAA negative subgroups (two-sided t-test p = 0.01). Specifically, asyn SAA positive subjects displayed reduced thickness and lower subcortical volumes than predicted suggesting greater cortical thinning and volume reduction in the presence of the misfolded alpha-synuclein (Fig. 5c). Notably, we did not observe this difference when comparing the 46 subjects based on their disease status or the presence of *LRRK2* pathogenic variants (both two-sided t-tests p > 0.05), demonstrating that the difference is driven by asyn SAA status.

Further region-by-region inspection of the difference between estimated and measured structure in asyn SAA positive participants highlighted pronounced degeneration in the frontal and parietal regions (Fig. 5d). In particular, regions showing the greatest difference between estimated and measured structure included the left transverse temporal gyrus, left pars opercularis, right caudal anterior cingulate, and left caudal middle frontal gyrus. Conversely, a few regions, such as the right temporal pole and the subcortical regions, exhibited milder PD-related atrophy. Collectively, our results characterized the links of positive asyn SAA status with greater brain atrophy, suggesting a worse prognosis and accelerated disease progression. These findings underscored the potential of CSF asyn SAA as a biomarker for monitoring or early detection of more severe neurodegeneration in Parkinson’s disease patients.

## Discussion

Mutations in the *LRRK2* gene are associated with autosomal and sporadic PD. *LRRK2* is a kinase with a role in lysosomal function and mitophagy, both of which are implicated in the pathogenesis of PD, and mutations in *LRRK2* lead to impaired clearance of alpha-synuclein fibrils^21^. Moreover, LRRK2 PD is similar to sporadic PD in age of onset, clinical manifestations and response to therapy^2^. Therefore, a better understanding of LRRK2 PD may provide insights into parkinsonian neurodegeneration more broadly and reveal how genetic predisposition translates into measurable changes in brain structure and function. Only recently has it been possible to collect enough genetic and neuroimaging measurements to investigate the *LRRK2* pathogenic variants in patients and non-manifesting variants carriers. In this pattern-matching study, we examined the impact of the *LRRK2* pathogenic variants in individuals with and without a PD diagnosis. This type of analysis required careful navigation of demographic variability and subgroup overlaps, which could otherwise obscure the true effects of *LRRK2* pathogenic variants and PD diagnosis.

To navigate through these challenges, we initially identified a brain pattern that distinguished carefully matched healthy controls from sporadic PD patients. The extracted brain pattern replicated previous findings of PD-related atrophy, particularly in posterior cortical regions^20^. Building on this validated pattern, we then used it as a benchmark to investigate structural changes in PD patients with the *LRRK2* pathogenic variants. Our findings revealed that LRRK2 PD patients exhibit milder cortical thinning and subcortical volume loss compared to those with sporadic PD. Moreover, the pattern of cortical thinning and subcortical volume loss in LRRK2 PD differed from the previously identified PD-related brain atrophy and may represent a unique MRI signature. In particular, we observed reduced cortical thinning in posterior cortical areas, and relative preservation of subcortical nuclei volumes. Recent evidence also documents a unique brainstem MRI signature for LRRK2 PD subjects that differentiate them from both sPD and HC^22^. While neuronal loss in the substantia nigra pars compacta and locus coeruleus represents a hallmark of PD^23,24^, LRRK2 PD patients displayed preservation of locus coeruleus neuromelanin^22^. In addition, previous reports also documented significantly higher basal forebrain volumes for LRRK2 PD compared to sPD and as well as HC^25–27^. We here expand these findings with notable preservation of cortical tissue in the temporal and occipital lobes.

From a clinical standpoint, LRRK2 PD patients show milder cognitive symptoms in comparison to sPD^28^. We observed a strong positive relationship between the predicted brain-structure metrics and participants’ cognitive performance. Similarly, increased cortical thickness was previously associated with higher MoCA scores^29^. Therefore, structural changes may be key indicators of disease progression and cognitive decline in Parkinson’s disease^30^. Since our LRRK2 PD subjects displayed less cortical thinning, these results support previous evidence of comparatively better cognitive function in this subgroup. Collectively, the reduced susceptibility to brain changes, along with the different and slower neurodegeneration observed in LRRK2 PD, suggests that the variants may define a feature of a biologically distinct subtype of PD. Such insights could lead to refined models of both disease progression and therapeutic strategies^31^.

We also examined the influence of the *LRRK2* pathogenic variants in subjects without a PD diagnosis. The analysis revealed that NMC did not exhibit significant cortical atrophy compared to healthy controls, suggesting no structural signs of subclinical Parkinson’s disease. Nevertheless, there is previous evidence of differences between HC and LRRK2 NMC including increased volume in cuneus^32^ or smaller volume in the hippocampal region^33^ for LRRK2 NMC. Furthermore, LRRK2 NMC displayed significantly increased brain cholinergic activity measured by positron emission tomography^26^. We here also observed several notable differences between predicted and measured thickness in LRRK2 NMC. These observations thus call for more well-powered studies investigating whether asymptomatic subjects remain structurally unaffected despite the genetic risk.

Finally, we investigated the contribution of Lewy pathology to PD-related brain atrophy. Notably, over one-third of LRRK2 PD patients may not have evidence of α- synuclein aggregates^3,16^. Recent study demonstrated significantly lower functional connectivity in bilateral caudates, and lower mean fractional anisotropy along the left superior fronto-occipital fasciculus for LRRK2 PD-SAA+ compared to LRRK2 PD-SAA-^34^. Despite the absence of significant volumetric differences between asyn SAA+ and asyn SAA− participants, the volumetric differences from healthy controls were less pronounced in PD-SAA−, potentially indicating a milder neurodegenerative process. Our analysis highlighted that asyn SAA+ participants with Lewy body pathology had greater cortical thinning than those without. As such, asyn SAA+ patients may represent yet another subtype, marked by faster disease progression, more severe brain atrophy, and a higher risk of dementia. However, because the asyn SAA+ and asyn SAA- groups differed in age, sex, education, and disease duration, the propensity score algorithm could only match a small fraction of the participants. Therefore, larger-scale studies are needed to elucidate the underlying biology of *LRRK2*-parkinsonism especially in the cases lacking alpha-synuclein aggregates. In summary, our observations are consistent with emerging evidence that PD may in fact comprise multiple neurodegenerative syndromes, all exhibiting similar motor and non-motor features yet rooted in diverse biochemical and molecular pathologies^35^.

Currently, the heterogeneity and the small size of clinical samples present a significant roadblock in studying MRI signatures of subjects with *LRRK2* pathogenic variants, in particular, and other rare genetic mutations that increase the risk for PD, in general (e.g., PRKN, PINK1, GBA, or SNCA)^9^. Consequently, the findings obtained from studying rare genetic mutations may have limited generalizability^36,37^. In addition, this low statistical power pushed prior research into investigating a small number of brain regions selected based on *a priori* hypothesis. While studies are hunting for early imaging markers, the heterogeneous nature of the disease complicates such a task^38^. Our findings suggest that there may be subtypes of PD, where the genetic background (such as *LRRK2* variants) modulates the spatial distribution and extent of cortical involvement.

Specifically, PD caused by *LRRK2* pathogenic variants appears to be associated with slower neurodegenerative processes. Notably, the preservation of cortical thickness and subcortical volume in *LRRK2* subjects reported here also included nucleus accumbens and pallidum, regions connected to basal forebrain. Collectively, in conjunction with the evidence of relatively spared basal forebrain in LRRK2 PD mentioned above, these findings point to an intact cholinergic system in LRRK2 PD subjects, which support the hypothesis of *LRRK2*- associated hypercholinergic activity that functions as an adaptive response to neurodegenerative processes^25^. While further voxel-based morphometry analyses do not report voxels surviving corrected statistical thresholds due to low sample sizes, our bespoke strategy also reports thickness preservation in large parts of the temporal and occipital cortex. This finding strengthens the idea of protective mechanisms associated with changes in cholinergic signaling. In other words, relatively enhanced cholinergic activity in LRRK2 PD subjects might help counteract the neurodegenerative processes and preserve cognitive function. A deeper understanding of how compensatory cholinergic mechanisms unfold is crucial for designing more targeted interventions since PD patients with cholinergic compensation could require distinct therapeutic approaches.

It is well established that PD, like other major brain disorders, involves a complex mechanism influenced by sex, age, and potentially other social identity factors^19,39^. Nevertheless, studying rare genetic variants often relies on convenience samples, which tend to be imbalanced, statistically underpowered, and prone to bias. Consequently, disparities in demographic and clinical characteristics can obscure relationships between PD diagnosis and key features of interest, such as brain architecture^40^. In the presence of confounding influences (e.g., age, sex, and disease duration), observed differences in brain structure may primarily reflect systematic biases rather than genuine effects of the disease^41,42^. Since our deconfounding strategies appear insufficient, it is crucial to first establish that groups are comparable in terms of their population background variation to ensure valid comparisons^18^.

We employed a carefully matched subgroup analysis using propensity score matching^43^. Propensity score adjustment allowed us to ensure comparability between groups by balancing the distribution of biases and confounders, thereby simulating the random assignment of subjects typically seen in a randomized trial. This analytical step enabled us to attribute observed brain atrophy to variables of interest—namely, PD diagnosis, *LRRK2* variant, asyn SAA status—rather than to demographic biases. In addition, we accompanied the careful participant matching by a pattern-matching approach to quantify differences between subgroups. Comparing the expression of well-established patterns of PD-related atrophy in smaller subgroups maximized our statistical power by reducing the need for multiple comparisons across regions, thereby yielding more robust findings that reflect widespread neural changes. This strategy opened up a more holistic view onto disease- related changes and allowed for the detection of complex structural patterns that might be missed when focusing on individual regions. These innovations provide a recipe for future studies to more accurately capture and interpret structural brain changes in complex brain disorders.

In conclusion, our pattern-matching strategy combined with careful modeling of individual heterogeneity revealed distinct MRI signatures among subgroups of PD patients. While the *LRRK2* pathogenic variants may be associated with a milder form of PD, Lewy body presence may lead to a more aggressive form of the disease. These genetic-imaging findings underscore the existence of distinct PD subtypes each shaped by genetic influences and linked to specific clinical and imaging features. Since *LRRK2* pathogenic variants are notably prevalent in certain populations and *LRRK2* gene presents a promising therapeutic target, these insights could facilitate the development of more precise prognostic models and personalized treatment strategies.

## Methods

### Participant cohort

Brain-imaging and clinical measurements were acquired as part of the Parkinson Progression Marker Initiative (PPMI). PPMI is a global landmark study, launched in 2010, aimed at identifying biomarkers to improve the diagnosis and progression tracking of Parkinson’s disease^17^ (https://www.ppmi-info.org). Each participating study site received approval from their local ethics committee and informed consent was obtained from all participants prior to enrolment in the study. PD patients and healthy controls met inclusion criteria as outlined by the PPMI.

From that resource, we have considered four subgroups with available MRI in our cross-sectional analysis: sporadic PD (sPD), PD patients with *LRRK2* pathogenic variants (LRRK2 PD), *LRRK2* non-manifesting carriers (LRRK2 NMC), and healthy controls (HC). Details on the genotyping methods used in PPMI are described elsewhere^44^. The sPD subgroup consisted of 288 PD patients (age 63.7±8.9 y.o., 178 males) without a known pathogenic mutation associated with PD (e.g., *LRRK2*, *GBA1*, *SNCA*, *Parkin*, *PINK1*). These patients were recruited to the study within 2 years of diagnosis and were not treated with dopaminergic medications during the first 6 months of enrolment. A subset of sPD patients participated in follow-up visits spanning 4 years, and their most recent imaging data was utilized for the analysis. The LRRK2 PD subgroup enrolled 76 *LRRK2* pathogenic variants carriers (age 65.3 ± 8.2 y.o., 43 males) within 7 years of PD diagnosis and irrespective of dopaminergic treatment status. Patients with mutations in both *LRRK2* and *GBA1* genes were excluded. The LRRK2 NMC subgroup consisted of 94 *LRRK2* pathogenic variants carriers without a PD diagnosis (age 62.0 ± 6.9 y.o., 40 males). Finally, 133 healthy controls (age 62.3 ± 10.2 y.o., 88 males) were absent of both pathogenic mutations and neurological diseases.

All participants with a diagnosis of PD had evidence of abnormal dopamine transporter imaging on single-photon emission computed tomography imaging^16,17^. The LRRK2 NMC were specifically recruited as first-degree relatives of PD patients of Ashkenazi Jewish origin from participating PPMI centres^45^. Most common pathogenic variant of *LRRK2 was* G2019S followed by R1441G^16^.

All demographic information can be found in Tables 1 and 2.

### Image acquisition and processing

All participants underwent T1-weighted MRI that followed standardized procedures and acquisition parameters (https://www.ppmi-info.org/study-design/research-documents-and-sops/). Surface-based cortical and volumetric subcortical processing was performed with FreeSurfer version 7.2^46^ to estimate vertex-wise maps of cortical thickness and subcortical volume. These two measures, as opposed to cortical surface area, have been shown to be most sensitive to neurodegeneration in PD^20^. Resulting brain maps were visually inspected for quality control and then parcellated into 68 cortical parcels according to the Desikan-Killiany atlas^47^ and 14 subcortical parcels according to Harvard-Oxford atlas^48^. These region of interest measures were computed by averaging the values of vertices within each parcel. Given that subcortical volumes scale with head size, models that examined these measures additionally included total intracranial volume as a covariate^20^.

### Alpha-synuclein seed amplification assay

Alpha-synuclein seed amplification assay (asyn SAA) was used to detect the presence of pathogenic alpha-synuclein aggregates in the cerebrospinal fluid (CSF) at the first visit of each participant. Details on the asyn SAA methodology and protocol used in PPMI have been previously described^12,49^. Each participant’s CSF sample was tested three separate times. A positive or negative asyn SAA was determined based on consensus, or lack thereof, across the three replicates. As a result, the asyn SAA provides a unique opportunity in our study to link alpha-synuclein aggregation to the structural brain changes captured through MRI.

### Continuous subgroup matching via propensity score calculation

To prepare the ground for dissecting the complexities of the dataset, we isolated comparable groups of healthy controls (i.e., without PD diagnosis and *LRRK2* pathogenic variants) and sporadic PD patients (e.g., with PD diagnosis and without *LRRK2* pathogenic variants). One of the most commonly used techniques for comparing participant groups is manually aligning participants based on age and sex. Balancing out subject groups, based on their population background variation ensures comparability between the groups under study and reduces potential confounding^18,19^. However, this manual approach becomes increasingly impractical with every additional covariate of undesired variation that needs to be considered. Therefore, we implemented a more flexible propensity score framework that natively aggregates a potentially large number of heterogeneous person characteristics into a single balancing index^40^.

Propensity scores were calculated using a logistic regression model, where the disease status was regressed on sex (encoded as a binary variable: male = 0, female = 1), age (in years), and site (encoded as dummy variables for each site location). The logistic regression model yielded a propensity score for each participant, representing the probability of carrying the diagnosis given the covariates:

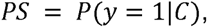

where y is the PD diagnosis, while C denotes a collection of covariates (e.g., sex, age, and site) to be accounted for.

Importantly, these propensity scores were based solely on the participant characteristics, that is, without access to input variables of primary scientific interest (i.e., brain region morphometry). Essentially, the propensity score distills each individual’s diversity characteristics into one number, facilitating more accurate and balanced group comparisons.

### Hungarian algorithm to align the propensity indexed participants into subgroups

After obtaining propensity scores for each HC and sPD participant through a logistic regression model, we proceeded to pair participants between the two groups under study using the Hungarian algorithm^50^. This optimization technique is designed to maximize the total similarity of pairing participants between the two groups (HC and sPD). We first constructed a cost matrix, where each entry represented the difference in propensity scores between a given sPD participant and a HC participant at hand. The Hungarian algorithm was then applied to this matrix to identify the optimal one-to-one matching, ensuring that the sum of differences in propensity scores across all matched pairs was minimized. Notably, in this matching process, we incorporated a caliper parameter to ensure that participants were only matched if their propensity scores were sufficiently close^40^. This approach modifies the cost matrix by assigning a prohibitively high cost to pairs exceeding the caliper checkpoint, ensuring that only those participants within the acceptable range are matched by pairing, thus improving the comparability and accuracy of the resulting samples (more details in Sup. Fig. 6). As an important consequence, since not all participants had sufficiently close propensity scores to be matchable, not all participants were selected for the following analysis (cf. corresponding stacked bar plots in figures). Instead, we have selected only closely matching pairs of participants, ensuring that the differences in propensity scores were within the acceptable range defined by the caliper. By aligning participants according to their propensity scores, the matching process created two subgroup samples (sPD and HC) that were balanced with respect to background variation in the population. In other words, the selected pairs of sPD and HC participants displayed similarly distributed covariates, reducing potential confounding effects and improving the comparability of the groups for subsequent analysis.

### Linear model to quantify structural changes in PD

After careful selection of matching pairs of HC and sPD subjects, we specified and estimated a series of linear regression models to quantify brain changes occurring in PD. Specifically, we estimated cortical thickness for cortical regions and volume for subcortical regions using PD diagnosis as model outcome (encoded as a binary variable: without diagnosis = 0, PD diagnosis = 1), sex (encoded as a binary variable: male = 0, female = 1), age (in years), and site (encoded as indicator variables, one for each site location). Models directed at subcortical volume used total intracranial volume as an additional input covariate following previous research^20^. The linear regression model follows a specification that can be formally expressed as:

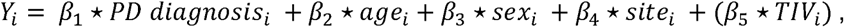

where Y_L_ corresponds to the regional morphometry of participant i and TJV is total intracranial volume. The analysis focused on the beta coefficient for PD diagnosis, with other covariates included to adjust for external sources of variation and ensure an accurate estimation of the effect of PD diagnosis on cortical thickness or subcortical volume.

Overall, we estimated 68 separate linear models corresponding to 68 cortical regions defined by the Desikan-Killiany cortical parcellation^47^ and 16 separate linear models corresponding to 16 regions defined by Harvard-Oxford subcortical atlas. These 84 total models allowed us to assess the observed structural brain changes associated with PD.

### Transferring the derived PD brain model to unseen subgroups

To further explore the relationships between brain morphometry, PD, and the *LRRK2* pathogenic variants, we utilized 68 linear models originally dedicated to the comparison between sPD and HC. We carried over these already estimated models—developed to quantify PD-related brain atrophy by estimating cortical thickness and subcortical volumes— to the other subgroups within our study. In other words, we leveraged these pre-trained models capturing PD-related brain atrophy patterns to explore differences among LRRK2 subgroups with distinct disease characteristics and demographics. This is because the sample sizes in these subgroups were too small at times to estimate a model afresh. Specifically, we focused on three group contrasts: LRRK2 PD versus sPD, LRRK2 NMC versus HC, and asyn SAA positive versus SAA negative participants. In short, our approach makes the quantitative analysis of these under-sampled subgroups possible in the first place.

As described (cf. above), the first step in these more nuanced subgroups involved creating matched groups of participant subsets based on a set of population background characteristics. Hence, we again employed propensity score matching to balance the groups in a way that reduces the effect of relevant sources of background variation in the population. For the comparison of LRRK2 PD and sPD participants, variant status was regressed on sex, age, disease duration, and site. The same set of covariates, with asyn SAA status as the outcome, was used to compare asyn SAA positive and negative participants. Finally, variant status was regressed on sex, age, and site in the HC versus LRRK2 NMC comparison (Fig. Sup. 5). Using the Hungarian algorithm to create pairs of participants matched on propensity scores minimized potential confounding and thus allowed for cleaner comparisons of brain morphometry across the different subgroups.

In the second step, we leveraged the robustness of the models trained on the larger sPD vs. HC comparison to generate reliable morphometric estimates across different subgroups. Carrying over the models, trained in earlier steps, in the new subgroups avoided the necessity of estimating new model parameters and thus addressed the low statistical power due to the small sample size. Consequently, we derived regional morphometry estimates for each matched participant in every participant from each subgroup.

### Testing morphometric predictions in data-poor subgroups at the whole brain level

Estimating regional morphometry allowed us to compare these estimates against actual measures of regional morphometry. In other words, we assessed the discrepancies between predicted and observed brain structure changes within each subgroup. Specifically, in each subgroup, we calculated the difference between predicted and measured morphometry for each participant. We then averaged these differences across participants to obtain a single aggregate estimate of predicted tissue differences for each brain region in each subgroup. Finally, we used paired t-tests to determine whether the differences between predicted and observed morphometry were statistically significant across regions, providing insight into the extent of brain structure changes associated with each subgroup at the whole brain level.

### Linking PD severity, cognition, and genetic risk to brain changes

To corroborate and further attach meaning to the brain morphometry estimates, we analyzed their association with several key characteristics routinely assessed in PD patients. Namely, we used the MDS-UPDRS part III, which is a comprehensive tool used to measure the severity of Parkinson’s disease. Here employed Part III of the UPDRS, which assesses motor manifestations, such as tremor, rigidity, and bradykinesia^51^. Furthermore, we also characterized every PD patient using the Montreal Cognitive Assessment (MoCA) screening tool designed to assess cognitive function and detect mild cognitive impairment. It evaluates various cognitive domains, including memory, attention, language, and executive functions^52^. Finally, we also used polygenic risk scores (PRS) for Parkinson’s disease computed by the PPMI consortium for every patient. PRS quantify an individual’s genetic predisposition to the disease by aggregating the effects of multiple genetic variants associated with increased risk. The PD PRS has been shown to be increased for LRRK2 PD^44^, suggesting that it may amplify the disease process in *LRRK2* pathogenic variants carriers. The PD PRS was computed and provided by the PPMI consortium from variants previously identified through genome-wide association studies^53^. We here used the modified PRS with LRRK2 locus excluded.

We collected UPDRS III and MoCA scores for every selected subject from the sPD versus LRRK2 PD comparison. We then used Pearson’s correlation to quantify the linear association strength between these motor and cognitive test scores and region-wise estimates of brain morphometry across participants. To complement this detailed examination and provide more general description, we also correlated UPDRS III and MoCA scores with morphometry estimates averaged across all regions. We employed the same strategy with PD PRS, but this time we used partial correlation analysis, controlling for the first 10 genetic principal components (PCs) to account for population structure and minimize potential confounding effects.

## Data Availability

Data used in the preparation of this article was obtained in May 2023 from the Parkinson's Progression Markers Initiative (PPMI) database (www.ppmi-info.org/access-data-specimens/download-data), RRID:SCR_006431. For up-to-date information on the study, visit www.ppmi-info.org.

## Acknowledgment

PPMI – a public-private partnership – is funded by the Michael J. Fox Foundation for Parkinson’s Research and funding partners, including 4D Pharma, Abbvie, AcureX, Allergan, Amathus Therapeutics, Aligning Science Across Parkinson’s, AskBio, Avid Radiopharmaceuticals, BIAL, BioArctic, Biogen, Biohaven, BioLegend, BlueRock Therapeutics, Bristol-Myers Squibb, Calico Labs, Capsida Biotherapeutics, Celgene, Cerevel Therapeutics, Coave Therapeutics, DaCapo Brainscience, Denali, Edmond J. Safra Foundation, Eli Lilly, Gain Therapeutics, GE HealthCare, Genentech, GSK, Golub Capital, Handl Therapeutics, Insitro, Jazz Pharmaceuticals, Johnson & Johnson Innovative Medicine, Lundbeck, Merck, Meso Scale Discovery, Mission Therapeutics, Neurocrine Biosciences, Neuron23, Neuropore, Pfizer, Piramal, Prevail Therapeutics, Roche, Sanofi, Servier, Sun Pharma Advanced Research Company, Takeda, Teva, UCB, Vanqua Bio, Verily, Voyager.

DB was supported by the Brain Canada Foundation, through the Canada Brain Research Fund, with the financial support of Health Canada, National Institutes of Health (NIH R01 AG068563A, NIH R01 DA053301-01A1, NIH R01 MH129858-01A1), the Canadian Institute of Health Research (CIHR 438531, CIHR 470425), the Healthy Brains Healthy Lives initiative (Canada First Research Excellence fund), the IVADO R3AI initiative (Canada First Research Excellence fund), and by the CIFAR Artificial Intelligence Chairs program (Canada Institute for Advanced Research). The funders had no role in study design, data collection and analysis, the decision to publish or the preparation of the manuscript.

## Code availability

The processing scripts and custom analysis software used in this work are available in a publicly accessible GitHub repository, along with examples of key visualizations in the paper: https://github.com/jakubkopal/LRRK2-MRI.

