## Supplementary material for "Carriers of *LRRK2* pathogenic variants show a milder, anatomically distinct brain signature of Parkinson’s disease": Sup. Fig.

### Supplementary Figures

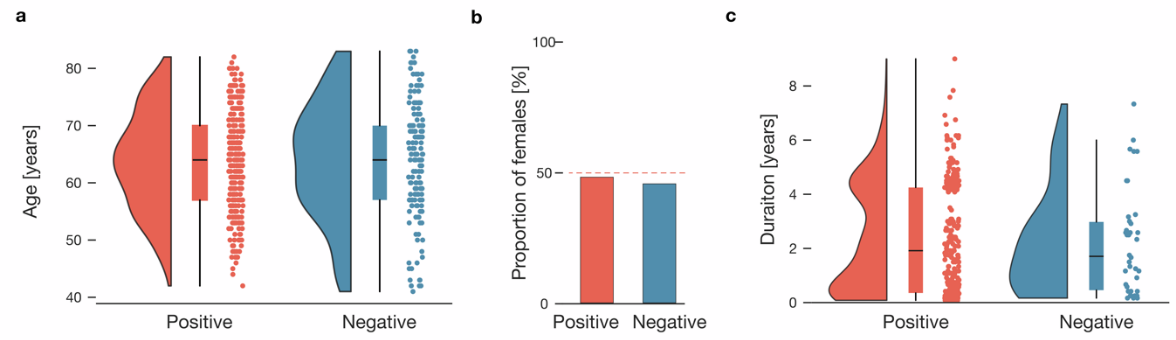


**Supplementary Figure 1: Key population characteristics differ based on SAA status**
**a.** SAA status and age distribution. Raincloud plots display the age of each participant in one of the two subgroups. **b.** SAA positive more prevalent in females. The red dotted line marks an equal ratio of male and female participants in a given subgroup. **c.** Differences in disease duration based on SAA status. Disease duration is plotted for all patients separated by the status of SAA test. Key demographic metrics, such as sex and disease duration, differe in groups of SAA positive and SAA negative participants.

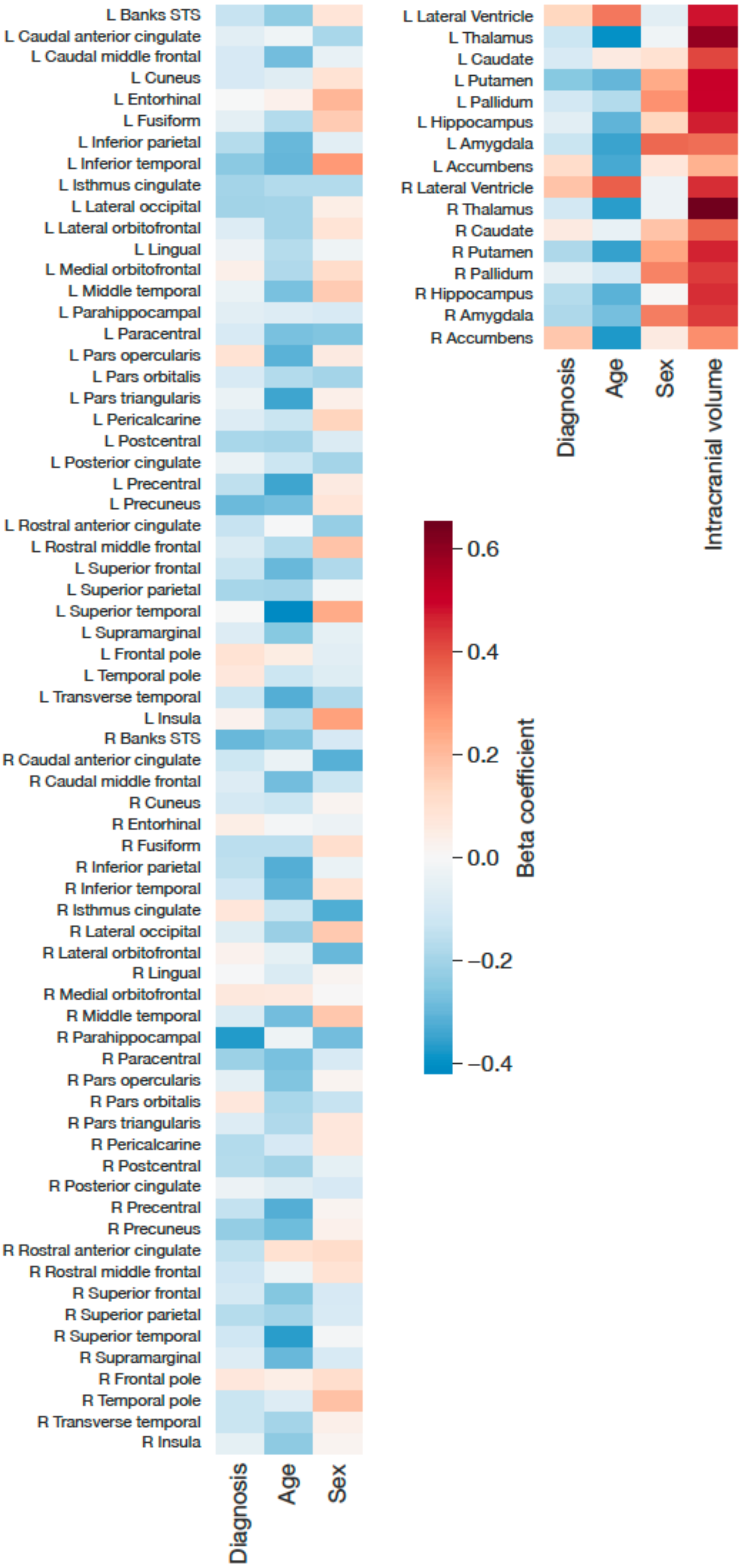

L Banks STS
L Caudal anterior cingulate
L Caudal middle frontal
L Cuneus
L Entorhinal
L Fusiform
L Inferior parietal
L Inferior temporal
L Isthmus cingulate
L Lateral occipital
L Lateral orbitofrontal
L Lingual
L Medial orbitofrontal
L Middle temporal
L Parahippocampal
L Paracentral
L Pars opercularis
L Pars orbitalis
L Pars triangularis
L Pericalcarine
L Postcentral
L Posterior cingulate
L Precentral
L Precuneus
L Rostral anterior cingulate
L Rostral middle frontal
L Superior frontal
L Superior parietal
L Superior temporal
L Supramarginal
L Frontal pole
L Temporal pole
L Transverse temporal
L Insula
R Banks STS
R Caudal anterior cingulate
R Caudal middle frontal
R Cuneus
R Entorhinal
R Fusiform
R Inferior parietal
R Inferior temporal
R Isthmus cingulate
R Lateral occipital
R Lateral orbitofrontal
R Lingual
R Medial orbitofrontal
R Middle temporal
R Parahippocampal
R Paracentral
R Pars opercularis
R Pars orbitalis
R Pars triangularis
R Pericalcarine
R Postcentral
R Posterior cingulate
R Precentral
R Precuneus
R Rostral anterior cingulate
R Rostral middle frontal
R Superior frontal
R Superior parietal
R Superior temporal
R Supramarginal
R Frontal pole
R Temporal pole
R Transverse temporal
R Insula
Diagnosis
Age
Sex
L Lateral Ventricle
L Thalamus
L Caudate
L Putamen
L Pallidum
L Hippocampus
L Amygdala
L Accumbens
R Lateral Ventricle
R Thalamus
R Caudate
R Putamen
R Pallidum
R Hippocampus
R Amygdala
R Accumbens
Diagnosis
Age
Sex
Intracranial volume
0.6
0.4
0.2
0.0
−0.2
−0.4
Beta coefficient

**Supplementary Figure 2: Role of disease diagnosis, age, and sex in determining Parkinson's disease outcomes**

Factors predicting Parkinson's disease. The heatmap displays beta coefficients of disease diagnosis, age, and sex from 68 linear models estimating cortical morphometry across 68 brain regions. For models estimating subcortical volume, effect size corresponding to total intracranial volume is displayed as well. Each cell represents the strength and direction of the association between the predictor (disease, age, sex, or intracranial volume) and the morphometric measure in a specific brain region. We predominantly observe negative effects of diagnosis and age, indicating that Parkinson's disease diagnosis is associated with brain atrophy, and this effect is exacerbated as age increases.

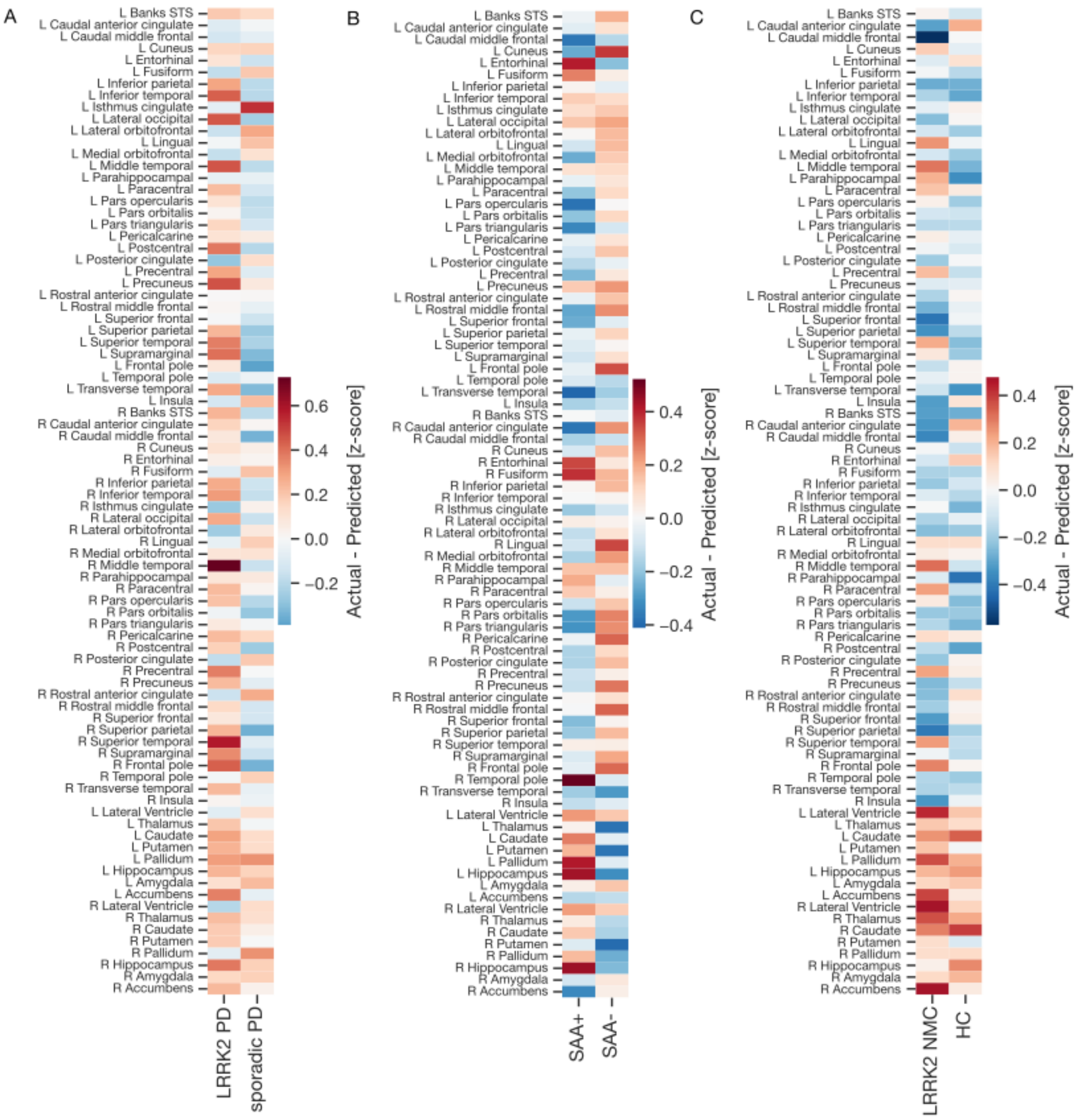


**Supplementary Figure 3: PD-model predictions across participant subgroups**

In each analytic scenario, we compare region-wise differences of measured and estimated morphometry between two subgroups of participants. Positive outcomes suggest preservation of brain structure while negative outcomes point towards pronounced degeneration. The heatmap displays region-wise differences for LRRK2 PD vs. sporadic PD comparison (a), SAA+ vs. SAA- comparison (b), and LRRK2 NMC vs. HC comparison (c).

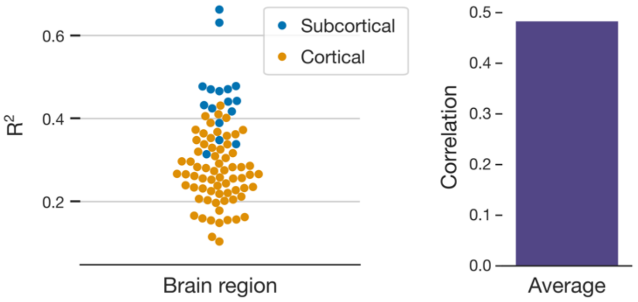


**Supplementary Figure 4: Linear models reliably capture brain morphometry**
**a.** Quantification of model reliability. We estimate 68 region-specific linear models predicting cortical thickness or subcortical volume using disease diagnosis, age, site, and, in the case of subcortical volume, total intracranial volume. The plot illustrates the proportion of variance explained by each model. **b.** Average prediction performance. To provide a global metric of performance across the 68 linear models, we calculate Pearson's correlation between the estimated and measured morphometry. The bar plot depicts the average correlation across all regions. The model fit analysis confirmed that the parameters were successful in accurately reflecting brain structure.

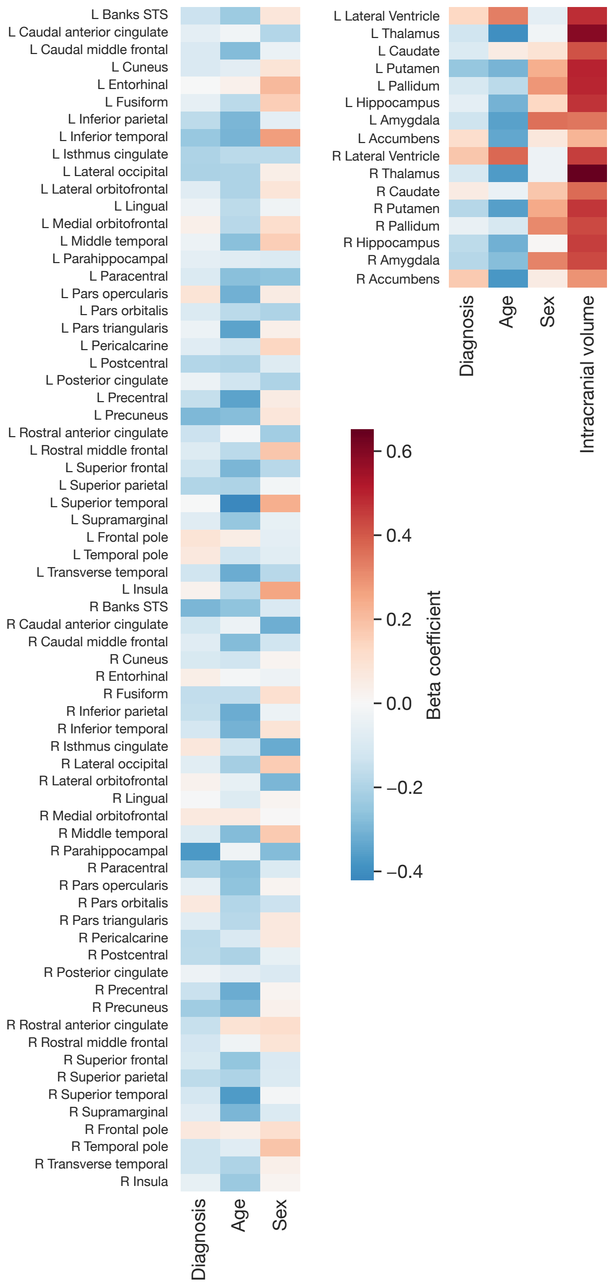


**Supplementary Figure 4: Role of disease diagnosis, age, and sex in determining Parkinson's disease outcomes**

Factors predicting Parkinson’s disease. The heatmap displays beta coefficients of disease diagnosis, age, and sex from 68 linear models estimating cortical morphometry across 68 brain regions. For models estimating subcortical volume, effect size corresponding to total intracranial volume is displayed as well. Each cell represents the strength and direction of the association between the predictor (disease, age, sex, or intracranial volume) and the morphometric measure in a specific brain region. We predominantly observe negative effects of diagnosis and age, indicating that Parkinson's disease diagnosis is associated with brain atrophy, and this effect is exacerbated as age increases.

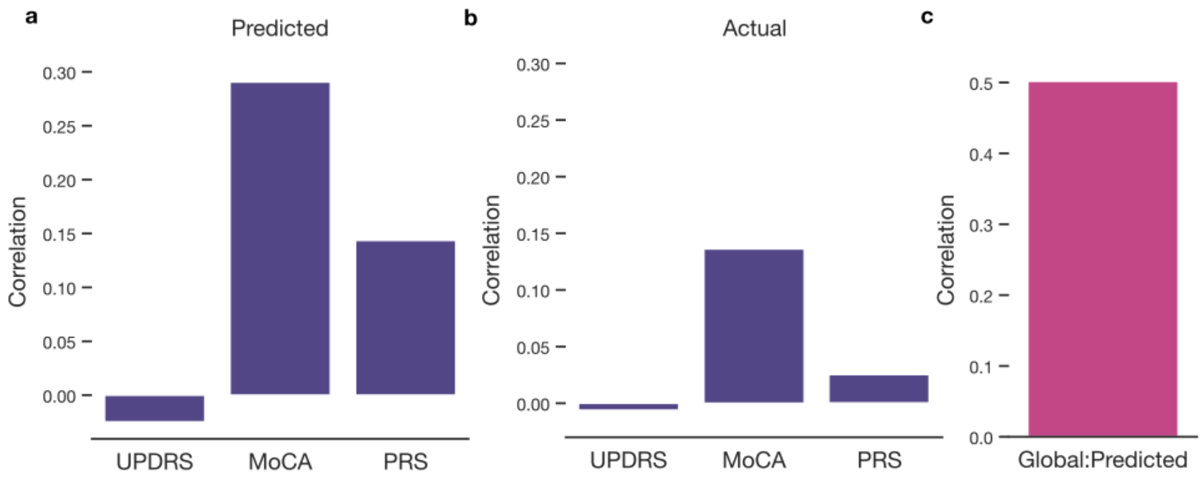


**Supplementary Figure 5: Zooming in on the relationships between patient characteristics and morphometry**

**a.** Predicted morphometry displays strong associations with patient characteristics. We examined how the Unified Parkinson's Disease Rating Scale (UPDRS), Montreal Cognitive Assessment (MoCA), and polygenic risk scores (PRS) for Parkinson’s disease correlate with both predicted brain morphometry across all sPD and LRRK2 participants. **b.** Associations with measured morphometry. We supplement the analyses by providing the associations between actual measured morphometry and patient characteristics. **c.** Association between estimated and predicted morphometry. Finally, we also calculated Person’s correlation between mean predicted and mean estimated morphometry across all regions. Overall, the associations of patient characteristics with estimated morphometry exceeded associations with actual measured structure.

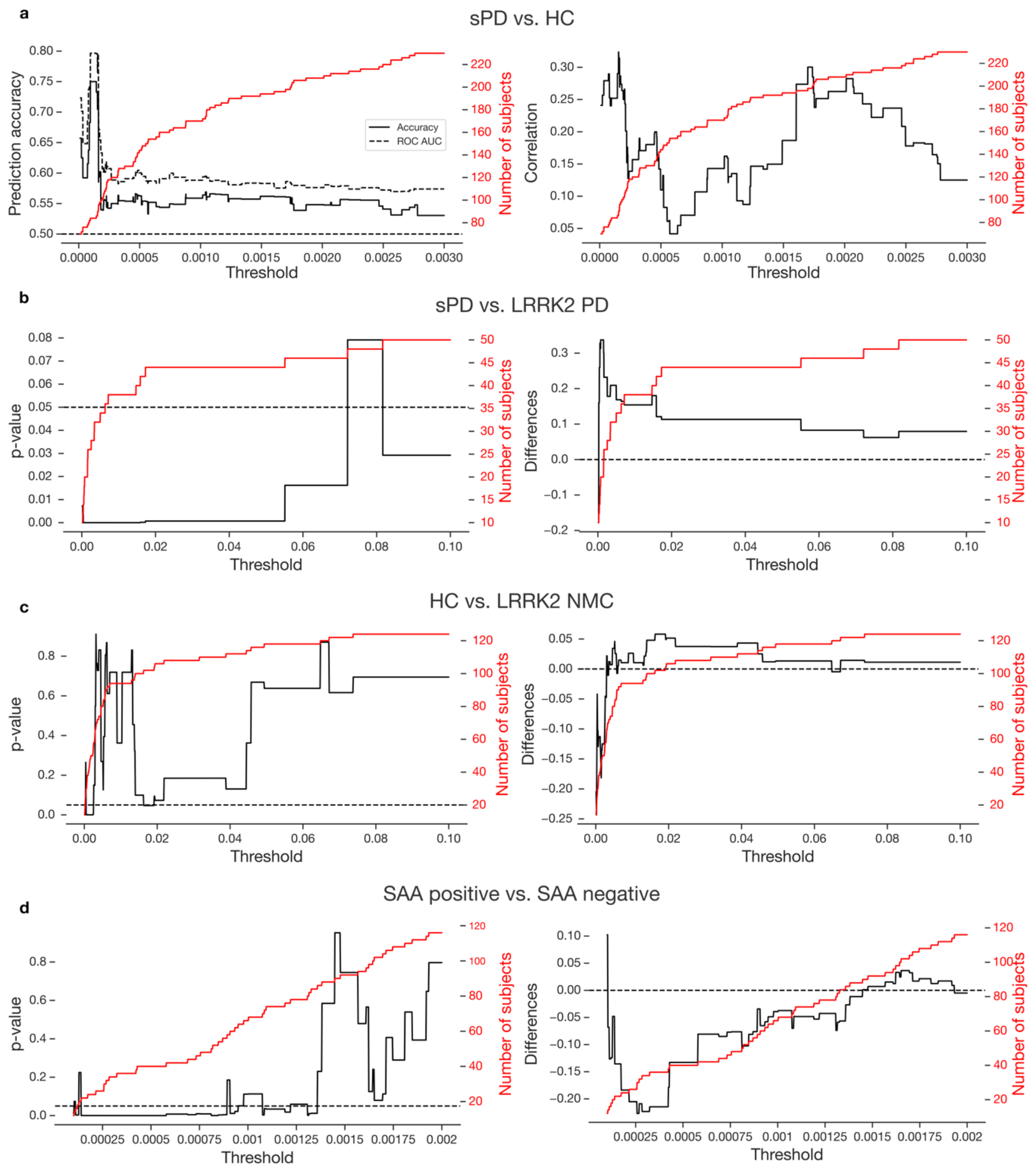


**Supplementary Figure 6: Optimization of caliper parameter in Hungarian algorithms**

In the Hungarian algorithm, a caliper is used to set a threshold for allowable differences in propensity scores, ensuring that participants are only matched if their scores fall within a predefined range, thereby improving the quality and comparability of the matched pairs. In other words, with a decreasing caliper, the matching process becomes more stringent, allowing only participants with very similar propensity scores to be paired, which enhances the precision of the matches but may reduce the number of eligible pairs. For each subgroup comparison, we investigated a range of caliper magnitudes in order to ensure the robustness of our findings. **a.** Model of PD-related atrophy. The left column depicts the relationship between caliper magnitude and prediction accuracy. The right column depicts the relationship between caliper magnitude and the similarity of obtained diagnosis effect sizes with those reported in Laansma et al., 2021. The red line depicts the number of selected participants. Caliper of 0.002 offered a large number of matched subjects with effects comparable to previous literature. **b.** The influence of *LRRK2* pathogenic variants in PD diagnosis. The left column depicts the relationship between caliper magnitude and p-value of group-wise differences. The right column depicts the relationship between caliper magnitude and the difference average between subgroups. The red line depicts the number of selected participants. Caliper of 0.01 offered a large number of matched subjects and prominent group differences. **c.** The influence of *LRRK2* pathogenic variants in subjects without PD diagnosis. The plots correspond to plots in **b** but for LRRK2 NMC and HC comparison. No caliper magnitude displayed consistent and robust group differences. **d.** The influence of SAA status. The plots correspond to plots in **b** but for SAA positive and SAA negative comparison. Caliper of 00075 offered a large number of matched subjects and prominent group differences. Overall, we carefully tuned the hyperparameters of our pipeline to ensure robust effects and minimize the influence of covariates.

### Supplementary Tables

**Supplementary Table 1: Regional effect sizes associated with Parkinson's disease**
The table contains beta coefficients of disease diagnosis, age, and sex from 68 linear models estimating cortical morphometry across 68 brain regions. For models estimating subcortical volume, effect size corresponding to total intracranial volume is displayed as well. Abbreviations: STS=superior temporal sulcus, L=left, R=right, ICV=intracranial volume.

| Brain region | ß diagnosis | ß sex | ß age | ß ICV |
|---|---|---|---|---|
| L Banks STS | -0,139 | 0,077 | -0,235 | |
| L Caudal anterior cingulate | -0,062 | -0,190 | -0,020 | |
| L Caudal middle frontal | -0,099 | -0,041 | -0,281 | |
| L Cuneus | -0,098 | 0,089 | -0,068 | |
| L Entorhinal | 0,001 | 0,207 | 0,033 | |
| L Fusiform | -0,059 | 0,159 | -0,174 | |
| L Inferior parietal | -0,167 | -0,062 | -0,294 | |
| L Inferior temporal | -0,243 | 0,272 | -0,300 | |
| L Isthmus cingulate | -0,198 | -0,177 | -0,171 | |
| L Lateral occipital | -0,204 | 0,045 | -0,199 | |
| L Lateral orbitofrontal | -0,082 | 0,083 | -0,198 | |
| L Lingual | -0,033 | -0,024 | -0,170 | |
| L Medial orbitofrontal | 0,035 | 0,111 | -0,180 | |
| L Middle temporal | -0,040 | 0,158 | -0,275 | |
| L Parahippocampal | -0,065 | -0,091 | -0,075 | |
| L Paracentral | -0,092 | -0,262 | -0,270 | |
| L Pars opercularis | 0,092 | 0,052 | -0,311 | |
| L Pars orbitalis | -0,092 | -0,197 | -0,178 | |
| L Pars triangularis | -0,040 | 0,035 | -0,343 | |
| L Pericalcarine | -0,075 | 0,143 | -0,135 | |
| L Postcentral | -0,190 | -0,084 | -0,200 | |
| L Posterior cingulate | -0,037 | -0,200 | -0,124 | |
| L Precentral | -0,153 | 0,052 | -0,343 | |
| L Precuneus | -0,289 | 0,080 | -0,276 | |
| L Rostral anterior cingulate | -0,138 | -0,224 | -0,005 | |
| L Rostral middle frontal | -0,087 | 0,181 | -0,171 | |
| L Superior frontal | -0,133 | -0,181 | -0,294 | |
| L Superior parietal | -0,195 | -0,017 | -0,199 | |
| L Superior temporal | 0,001 | 0,237 | -0,421 | |
| L Supramarginal | -0,081 | -0,055 | -0,249 | |
| L Frontal pole | 0,092 | -0,062 | 0,047 | |
| L Temporal pole | 0,068 | -0,073 | -0,127 | |
| L Transverse temporal | -0,127 | -0,180 | -0,321 | |
| L Insula | 0,028 | 0,253 | -0,175 | |
| R Banks STS | -0,300 | -0,099 | -0,261 | |

| | | | | |
|---|---|---|---|---|
| R Caudal anterior cingulate | -0,123 | -0,314 | -0,040 | |
| R Caudal middle frontal | -0,075 | -0,128 | -0,282 | |
| R Cuneus | -0,103 | 0,022 | -0,132 | |
| R Entorhinal | 0,047 | -0,029 | -0,011 | |
| R Fusiform | -0,159 | 0,103 | -0,161 | |
| R Inferior parietal | -0,151 | -0,038 | -0,318 | |
| R Inferior temporal | -0,115 | 0,092 | -0,308 | |
| R Isthmus cingulate | 0,075 | -0,327 | -0,133 | |
| R Lateral occipital | -0,071 | 0,164 | -0,219 | |
| R Lateral orbitofrontal | 0,029 | -0,296 | -0,053 | |
| R Lingual | -0,006 | 0,021 | -0,088 | |
| R Medial orbitofrontal | 0,063 | 0,005 | 0,057 | |
| R Middle temporal | -0,091 | 0,170 | -0,282 | |
| R Parahippocampal | -0,367 | -0,280 | -0,026 | |
| R Paracentral | -0,215 | -0,091 | -0,269 | |
| R Pars opercularis | -0,059 | 0,018 | -0,261 | |
| R Pars orbitalis | 0,068 | -0,138 | -0,188 | |
| R Pars triangularis | -0,076 | 0,068 | -0,181 | |
| R Pericalcarine | -0,175 | 0,067 | -0,098 | |
| R Postcentral | -0,167 | -0,051 | -0,204 | |
| R Posterior cingulate | -0,036 | -0,097 | -0,070 | |
| R Precentral | -0,143 | 0,015 | -0,321 | |
| R Precuneus | -0,232 | 0,034 | -0,283 | |
| R Rostral anterior cingulate | -0,147 | 0,114 | 0,097 | |
| R Rostral middle frontal | -0,118 | 0,087 | -0,028 | |
| R Superior frontal | -0,102 | -0,097 | -0,256 | |
| R Superior parietal | -0,170 | -0,093 | -0,201 | |
| R Superior temporal | -0,112 | -0,012 | -0,363 | |
| R Supramarginal | -0,081 | -0,094 | -0,298 | |
| R Frontal pole | 0,070 | 0,110 | 0,043 | |
| R Temporal pole | -0,134 | 0,182 | -0,080 | |
| R Transverse temporal | -0,135 | 0,039 | -0,201 | |
| R Insula | -0,055 | 0,017 | -0,237 | |
| L Lateral Ventricle | 0,146 | -0,068 | 0,357 | 0,514 |
| L Thalamus | -0,137 | -0,025 | -0,427 | 0,637 |
| L Caudate | -0,099 | 0,105 | 0,061 | 0,440 |
| L Putamen | -0,265 | 0,255 | -0,324 | 0,537 |
| L Pallidum | -0,115 | 0,308 | -0,186 | 0,528 |
| L Hippocampus | -0,069 | 0,144 | -0,329 | 0,504 |
| L Amygdala | -0,145 | 0,384 | -0,377 | 0,377 |
| L Accumbens | 0,124 | 0,083 | -0,361 | 0,236 |
| R Lateral Ventricle | 0,196 | -0,034 | 0,398 | 0,483 |
| R Thalamus | -0,116 | -0,032 | -0,394 | 0,699 |

| | | | | |
|---|---|---|---|---|
| R Caudate | 0,060 | 0,191 | -0,048 | 0,395 |
| R Putamen | -0,196 | 0,265 | -0,379 | 0,497 |
| R Pallidum | -0,053 | 0,339 | -0,117 | 0,460 |
| R Hippocampus | -0,184 | 0,010 | -0,339 | 0,482 |
| R Amygdala | -0,198 | 0,346 | -0,295 | 0,461 |
| R Accumbens | 0,184 | 0,060 | -0,404 | 0,314 |

**Supplementary Table 2: The relationship between patient characteristics and predicted morphometry**

The table depicts the linear association of Unified Parkinson's Disease Rating Scale (UPDRS), Montreal Cognitive Assessment (MoCA) scores, and polygenic risk scores (PRS) for Parkinson's disease with estimated morphometry separately for every brain region. The correlation with PRS was calculated as partial correlation while controlling for first 10 genetic principal components. Abbreviations: STS=superior temporal sulcus, L=left, R=right.

| Region | UPDRS | MoCA | PRS |
|---|---|---|---|
| L Banks STS | 0,06 | 0,24 | 0,13 |
| L Caudal anterior cingulate | -0,03 | 0,07 | 0,03 |
| L Caudal middle frontal | -0,15 | 0,27 | 0,13 |
| L Cuneus | 0,11 | 0,21 | 0,06 |
| L Entorhinal | -0,11 | -0,12 | -0,03 |
| L Fusiform | -0,05 | 0,11 | 0,07 |
| L Inferior parietal | 0,02 | 0,20 | -0,02 |
| L Inferior temporal | -0,10 | 0,21 | -0,06 |
| L Isthmus cingulate | -0,10 | 0,07 | 0,10 |
| L Lateral occipital | -0,01 | 0,14 | -0,01 |
| L Lateral orbitofrontal | -0,02 | 0,13 | -0,03 |
| L Lingual | -0,01 | 0,18 | 0,13 |
| L Medial orbitofrontal | 0,01 | 0,13 | -0,03 |
| L Middle temporal | 0,00 | 0,22 | 0,01 |
| L Parahippocampal | 0,02 | 0,00 | -0,01 |
| L Paracentral | -0,06 | 0,33 | 0,17 |
| L Pars opercularis | 0,01 | 0,25 | 0,07 |
| L Pars orbitalis | -0,12 | 0,12 | -0,05 |
| L Pars triangularis | 0,01 | 0,20 | 0,05 |
| L Pericalcarine | -0,02 | 0,08 | 0,10 |
| L Postcentral | -0,05 | 0,38 | 0,10 |
| L Posterior cingulate | 0,00 | 0,05 | 0,11 |
| L Precentral | -0,06 | 0,31 | 0,16 |
| L Precuneus | 0,10 | 0,27 | 0,09 |
| L Rostral anterior cingulate | 0,05 | 0,11 | -0,02 |
| L Rostral middle frontal | 0,06 | 0,13 | 0,04 |
| L Superior frontal | -0,03 | 0,21 | 0,12 |
| L Superior parietal | 0,04 | 0,21 | 0,05 |
| L Superior temporal | -0,11 | 0,30 | 0,10 |
| L Supramarginal | 0,01 | 0,22 | -0,02 |
| L Frontal pole | -0,09 | -0,14 | -0,08 |
| L Temporal pole | 0,03 | 0,10 | 0,09 |
| L Transverse temporal | -0,10 | 0,25 | 0,22 |
| L Insula | 0,00 | 0,10 | 0,12 |
| R Banks STS | -0,01 | 0,19 | 0,11 |

| | | | |
|---|---|---|---|
| R Caudal anterior cingulate | 0,07 | 0,07 | 0,07 |
| R Caudal middle frontal | -0,08 | 0,26 | 0,05 |
| R Cuneus | 0,11 | 0,21 | 0,04 |
| R Entorhinal | -0,13 | 0,13 | -0,05 |
| R Fusiform | -0,04 | 0,12 | 0,05 |
| R Inferior parietal | 0,02 | 0,29 | -0,02 |
| R Inferior temporal | -0,02 | 0,22 | -0,04 |
| R Isthmus cingulate | -0,07 | 0,02 | 0,10 |
| R Lateral occipital | 0,05 | 0,25 | 0,06 |
| R Lateral orbitofrontal | 0,03 | 0,17 | -0,07 |
| R Lingual | 0,01 | 0,24 | 0,04 |
| R Medial orbitofrontal | 0,14 | -0,14 | -0,08 |
| R Middle temporal | 0,03 | 0,24 | -0,07 |
| R Parahippocampal | -0,09 | 0,02 | 0,06 |
| R Paracentral | -0,06 | 0,33 | 0,13 |
| R Pars opercularis | 0,07 | 0,23 | -0,04 |
| R Pars orbitalis | 0,00 | 0,14 | -0,10 |
| R Pars triangularis | 0,06 | 0,08 | -0,09 |
| R Pericalcarine | 0,07 | 0,13 | 0,06 |
| R Postcentral | -0,04 | 0,26 | 0,08 |
| R Posterior cingulate | 0,03 | 0,08 | 0,08 |
| R Precentral | -0,06 | 0,31 | 0,14 |
| R Precuneus | 0,03 | 0,29 | 0,09 |
| R Rostral anterior cingulate | 0,14 | -0,26 | 0,00 |
| R Rostral middle frontal | 0,10 | -0,06 | -0,08 |
| R Superior frontal | 0,00 | 0,21 | 0,06 |
| R Superior parietal | 0,05 | 0,25 | 0,06 |
| R Superior temporal | -0,04 | 0,32 | -0,01 |
| R Supramarginal | 0,04 | 0,24 | -0,07 |
| R Frontal pole | 0,10 | -0,05 | -0,12 |
| R Temporal pole | -0,02 | 0,01 | -0,05 |
| R Transverse temporal | -0,06 | 0,25 | 0,22 |
| R Insula | -0,07 | 0,15 | 0,11 |
| L Lateral Ventricle | 0,09 | -0,16 | -0,16 |
| L Thalamus | -0,09 | 0,22 | 0,05 |
| L Caudate | 0,02 | 0,08 | 0,02 |
| L Putamen | -0,11 | 0,13 | 0,08 |
| L Pallidum | 0,06 | 0,14 | -0,07 |
| L Hippocampus | -0,08 | 0,19 | 0,01 |
| L Amygdala | 0,00 | 0,22 | 0,01 |
| L Accumbens | -0,09 | 0,26 | 0,03 |
| R Lateral Ventricle | 0,10 | -0,17 | -0,16 |
| R Thalamus | -0,03 | 0,24 | -0,01 |

| | | | |
|---|---|---|---|
| R Caudate | -0,04 | 0,11 | 0,06 |
| R Putamen | -0,12 | 0,12 | 0,12 |
| R Pallidum | 0,00 | 0,04 | 0,02 |
| R Hippocampus | -0,08 | 0,21 | 0,00 |
| R Amygdala | -0,02 | 0,13 | 0,03 |
| R Accumbens | -0,04 | 0,30 | 0,05 |
